# Joint Model with Random Changepoints for Longitudinal Measures and Semi-Competing Risks

**DOI:** 10.1101/2024.06.26.24309540

**Authors:** Xuzhi Wang, Yorghos Tripodis, Michael LaValley, Chunyu Liu

## Abstract

Dementia is characterized by a gradual loss of cognitive abilities, often accompanied by an accelerated decline preceding diagnosis. Cognitive decline in patients with dementia may lead to complications that impact mortality. Changepoint models have been proposed to identify when cognitive decline accelerates and how it progresses. Joint models with changepoints further account for dropout due to death or dementia. However, few joint models with changepoints consider semi-competing risks (e.g., dementia and death) by distinguishing transitions between various health states. We propose a joint model that combines a random changepoint model for cognitive decline with an illness-death model for semi-competing risks. We examine the proposed model with different changepoint formulations and association structures that connect the longitudinal and survival processes. Simulations demonstrate that our proposed model effectively estimates changepoints and characterizes the influence of longitudinal measures on transitions between health states. Real data application indicates associations of changepoints in cognitive trajectories with dementia and death in a community-based cohort. Our method provides insights into cognitive health in the presence of semi-competing risks.

## 1 Introduction

Dementia is a progressive condition that impairs memory, thinking, and decision-making, significantly affecting daily activities [1]. It is the leading cause of disability among the elderly [2]. Dementia begins with gradual cognitive decline, followed by an accelerated deterioration before diagnosis. Therefore, early detection of cognitive change is crucial for preventing and treating dementia. Various statistical models have been proposed to better understand the shape of cognitive decline over time, including mixed effects models with polynomial functions and changepoint models. Compared to polynomial function models, changepoint models help identify when cognitive decline accelerates and have been applied in various medical areas [3]. Hall et al. (2000) proposes a piecewise linear mixed model that assumes an abrupt change before and after the changepoint [3, 4]. To better reflect real-world scenarios, more flexible models with quadratic or polynomial transition functions have been proposed for smooth changes around the changepoint [5, 6]. Further extensions include handling bivariate longitudinal measures and multiple random changepoints [3, 7, 8]. Incorporating changepoint models into joint framework has led to models capable of handling informative dropout due to death or dementia and analyzing the association between cognitive decline and dementia [9-13]. Jacqmin-Gadda et al. (2006) proposes a joint model that integrates a piecewise mixed model with a random changepoint for cognitive decline and a log-normal model for time to dementia [9]. Subsequently, Yu et al. (2010) extends this approach by considering two competing risks: dementia and dementia-free death [10]. Tapsoba et al. (2011) focuses on the time-to-event process of the joint model, treating the longitudinal process as a time-varying covariate measured with error [12]. Furthermore, Wang (2021) proposes a joint model that includes a random changepoint model with a smooth change and competing risks [13].

However, there is limited literature on the joint models with a changepoint that account for semi-competing risks (e.g., dementia and death) by distinguishing transitions between health states, such as dementia, death following dementia, and death without dementia. Dantan et al. (2010) proposes a joint multistate model that includes a changepoint and investigates transition intensities between four health states [14]. However, they only consider one longitudinal outcome and assume identical transition intensities from three transient states to death. Rouanet et al. (2016) develops a joint model combining a mixed effects model, which includes a class-specific fixed changepoint, with an illness-death model [15]. However, this joint model uses a latent class approach without explicitly specifying the relationship between the changepoint and semi-competing risks. In our paper, the main objective is to estimate changepoints in longitudinal cognitive trajectories and examine their impact on transitions between health states. We propose a joint model for multiple longitudinal measures with changepoints and semi-competing risks. Our proposed joint model is decomposed into a multivariate random changepoint model for the longitudinal data and an illness-death model that estimates the transitions between health states for the semi-competing risks data. To demonstrate the flexibility of our proposed method for handling various data assumptions, we evaluate two formulations of changepoints, alongside two types of association structures connecting the longitudinal and survival processes.

The rest of the paper proceeds as follows. In Section 2, we present the proposed method. Section 3 describes the estimation of the proposed method. Section 4 evaluates the performance of our proposed method via a simulation study. Section 5 provides the simulation results. In Section 6, we apply our proposed approach to a cohort dataset. We summarize our main findings and future extensions in Section 7.

## 2 Methods

### 2.1 Random Changepoint model: Longitudinal Sub-model

In the longitudinal part of our proposed joint model, we consider two models with different changepoint formulations: the piecewise model with a sudden change around the changepoint and the bent-cable model with a gradual change.

#### Piecewise (PW) model

Let *Y*_*ki*_(*t*_*ij*_) be the value of the *k*^th^ longitudinal measure at time *t*_*ij*_ for subject *i*, where *i* = 1, 2, …, *N, k* = 1, 2, …, *K*, and *j* = 1,2, …, *n*_*ki*_. *N* denotes the total number of participants, and *K* denotes the total number of longitudinal measures. *n*_*ki*_ represents the number of repeated measurements of the *k*th longitudinal measure for participant *i*. Measurement times may differ among *K* longitudinal measures and across *N* participants. Let *Z*_*i*_ be a baseline covariate for participant *i*. The piecewise model can be formulated as

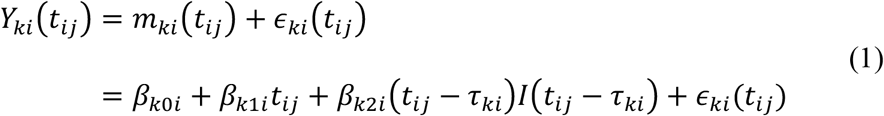

where *m*_*ki*_(*t*_*ij*_) denotes the true unobserved value of *Y*_*ki*_(*t*_*ij*_). The random error *ϵ*_*ki*_(*t*_*ij*_) is distributed as 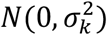 and is independent of all other parameters. *I*(·) denotes the indicator function such that *Y*_*ki*_(*t*_*ij*_) = *β*_*k*0*i*_ + *β*_*k*1*i*_*t*_*ij*_ + *ϵ*_*ki*_(*t*_*ij*_) when *t*_*ij*_ ≤ *τ*_*ki*_, and *Y*_*ki*_(*t*_*ij*_) = *β*_*k*0*i*_ + *β*_*k*1*i*_*t*_*ij*_ + *β*_*k*2*i*_(*t*_*ij*_ − *τ*_*ki*_) + *ϵ*_*ki*_(*t*_*ij*_) when *t*_*ij*_ > *τ*_*ki*_. Here, *τ*_*ki*_ is the random changepoint for subject *i* and longitudinal measure *k*. The random changepoint is treated as a latent variable and it may not always occur within a participant’s observed follow-up period. *β*_*k*0*i*_ represents the subject-specific intercept, *β*_*k*1*i*_ is the subject-specific slope before the changepoint *τ*_*ki*_, and *β*_*k*2*i*_ denotes the difference in slope before and after the changepoint *τ*_*ki*_.

Each of the *K* random changepoints *τ*_*ki*_ includes a population-level fixed effect, which may depend on a baseline covariate *Z*_*i*_, and an individual-level random effect *b*_*τki*_: *τ*_*ki*_ = *β*_*τk*0_ + *β*_*τk*1_*Z*_*i*_ + *b*_*τki*_. We assume the vector of individual-level random effects in changepoints across *K* longitudinal measures follows a multivariate normal distribution: (*b*_*τ*1*i*_, *b*_*τ*2*i*_, …, *b*_*τKi*_)^′^∼*MVN*(0, Σ_*τ*_). Similarly, the subject-specific intercepts and slopes in each of the *K* longitudinal measures can be decomposed into a fixed part and a random part, such that *β*_*kgi*_ = *β*_*kg*_ + *b*_*kgi*_, *g* = 0, 1, 2. Let ***b***_***ki***_ = (*b*_*k*0*i*_, *b*_*k*1*i*_, *b*_*k*2*i*_)′ for the *k*th longitudinal measure, then the vector of ***b***_***ki***_ across *K* longitudinal measures follows a multivariate normal distribution: (***b***_**1*i***_, ***b***_**2*i***_, …, ***b***_***Ki***_)′∼*MVN*(0, Σ_*b*_). Therefore, we assume all *K* longitudinal measures are correlated via the random changepoints, as well as through the random intercepts and slopes.

The piecewise model assumes an abrupt change around the changepoint and gives us a straightforward interpretation for the slope parameters [3, 16] (left panel of **Fig. 1**). However, the abrupt change in trajectory may not always align with real-world scenarios, where changes often occur more gradually, such as cognitive decline preceding dementia onset [8]. As an alternative, the bent-cable model assumes a smooth change between two linear phases around the changepoint [5].

**Fig. 1.**
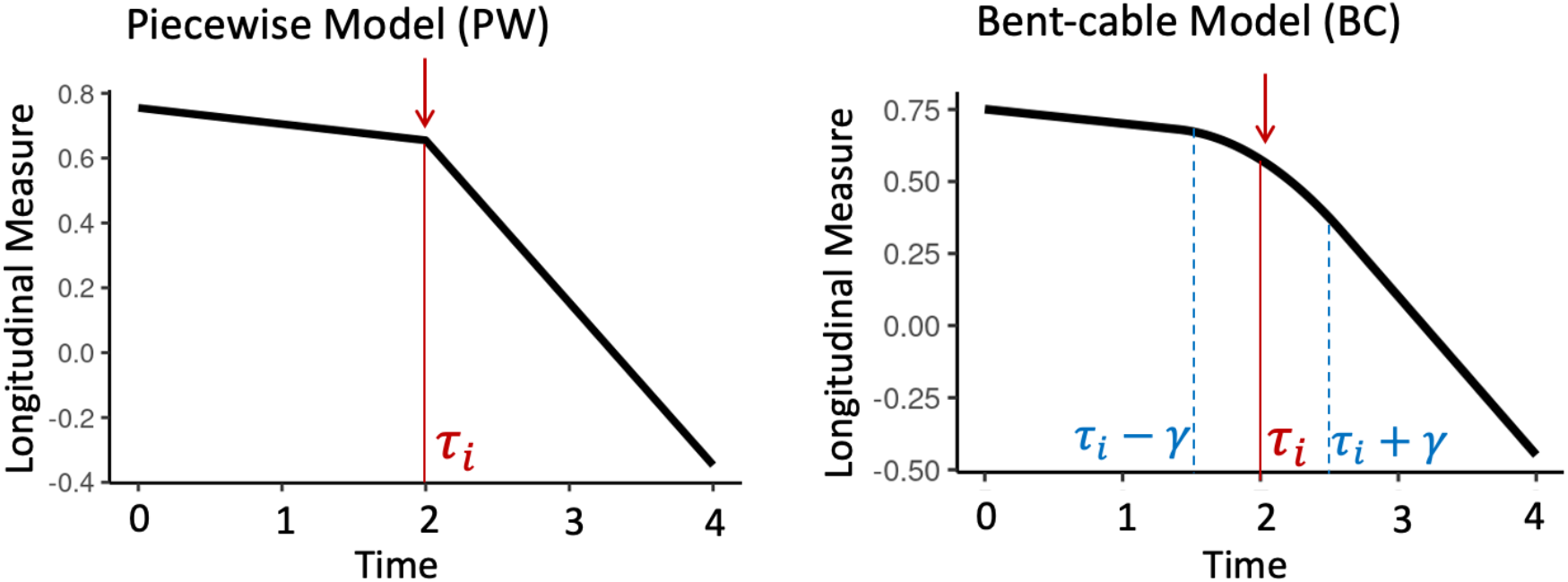
The visualization of the piecewise model (left) and the bent-cable model (right). The y-axis represents the value of a longitudinal measure, and the x-axis represents time. ***τ***_***i***_ is the changepoint for participant ***i***. [***τ***_***i***_ − ***γ, τ***_***i***_ + ***γ***] denotes the smooth change zone for the bent-cable model

#### Bent-cable (BC) model

Let *Y*_*ki*_(*t*_*ij*_) be the value of the *k*^th^ longitudinal measure at time *t*_*ij*_ for subject *i*, the model can be formulated as

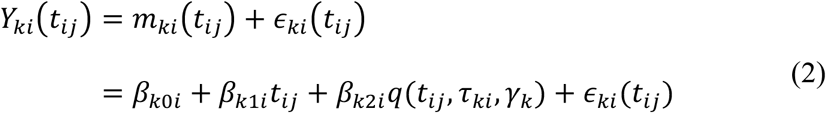

where the smooth function *q*(*t*_*ij*_, *τ*_*ki*_, *γ*_*k*_) is expressed as

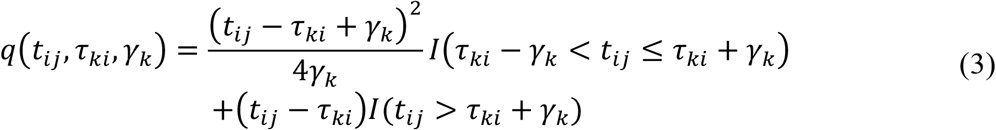

with *I*(·) as an indicator function. Using this function, we define the smooth change zone as the interval [*τ*_*ki*_ − *γ*_*k*_, *τ*_*ki*_ + *γ*_*k*_], where *γ*_*k*_ is the half width of the smooth change zone around the changepoint *τ*_*ki*_. All the other parameters share the same definitions as those in the piecewise model. When *γ*_*k*_ approaches 0, the smooth change zone shifts to an abrupt change at the changepoint, which closely approximates the bent-cable model to a piecewise model. The bent-cable model adds more flexibility to the piecewise model by defining a smooth change zone. Both the bent-cable model and the piecewise model share the same interpretation for the intercept, slopes and random changepoint, which facilitates model comparison. **Fig. 1** illustrates the visualization of both models, clearly depicting their respective features and differences.

### 2.2 Illness-death Model: Survival Sub-model

During follow-up in the dementia study, a participant may undergo one or more of the three health states: healthy state (state 0), dementia state (state 1), and death state (state 2) (**Fig. 2**). Since the occurrence of dementia (non-terminal event) is subject to death (terminal event), these events are considered as semi-competing risks [17]. Let *t*_1_ and *t*_2_ be the times to non-terminal and terminal events. In the survival part of our joint model, we use an illness-death model [18, 19]. The model is designed to capture the dynamics of transitioning between the three distinct health states via three hazard functions: *h*_1_(*t*_1_) for the hazard of dementia at time *t*_1_ given that neither dementia nor death occurred before time *t*_1_, *h*_2_(*t*_2_) denotes the hazard of death at time *t*_2_ given that neither events occurred before time *t*_2_, and *h*_3_(*t*_2_|*t*_1_) is the hazard of death at time *t*_2_ conditional on the occurrence of dementia at time *t*_1_. Specifically, we construct the following three parametric proportional hazards models:

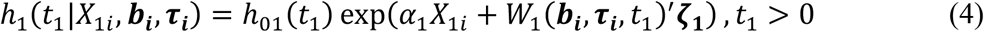

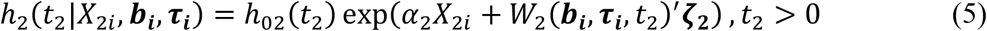

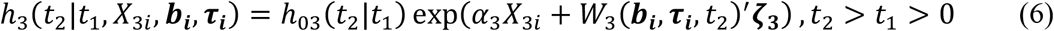

**Fig. 2.**
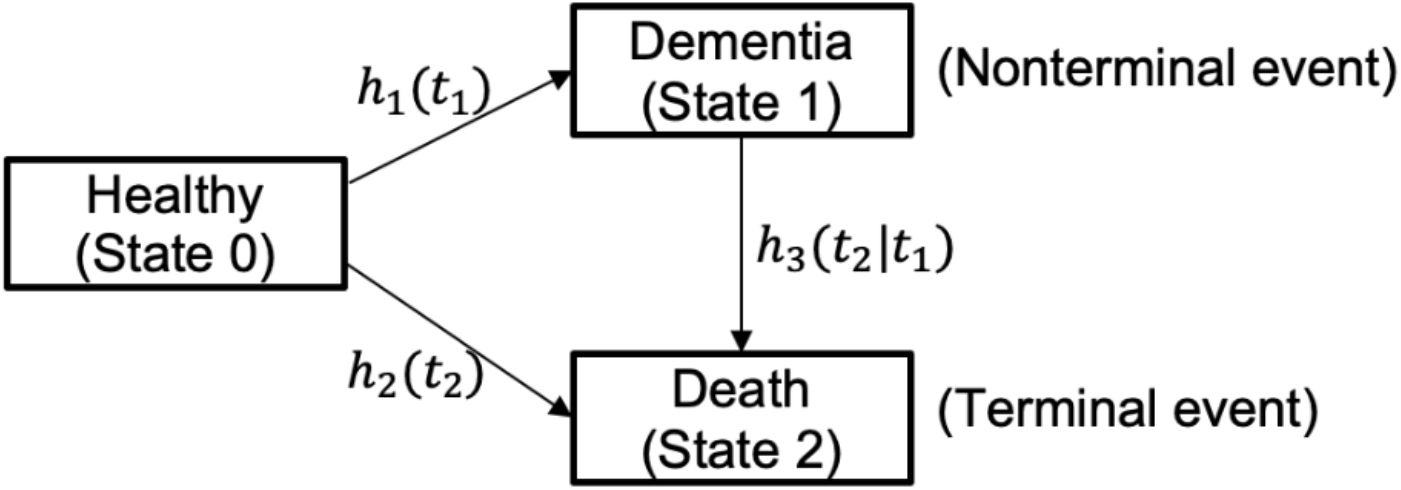
Three health states with semi-competing risks. ***h***_**1**_(***t***_**1**_) is the hazard function for dementia at time ***t***_**1**_, ***h***_**2**_(***t***_**2**_) is the hazard function for death at time ***t***_**2**_, and ***h***_**3**_(***t***_**2**_|***t***_**1**_) denotes the hazard function for death at time ***t***_**2**_ given that the participant develops dementia at time ***t***_**1**_

Let *g* ∈ {1,2,3} and *h*_0*g*_ is the transition-specific baseline hazards (Weibull or piecewise constant for example). *α*_*g*_ is the transition-specific log-hazard ratio regression parameters associated with the baseline covariates *X*_*gi*_. *W*_*g*_ is a transition-specific function of individual-level random effects (i.e., random intercepts and slopes ***b***_***i***_ and random changepoints ***τ***_***i***_) from the longitudinal sub-model, which defines the association structure between the longitudinal and survival sub-models. ***ζ***_***g***_ is a transition-specific vector of association coefficients for *W*_*g*_, which quantifies the impact of the longitudinal dynamics on the hazards. We further assume a semi-Markov model for the conditional baseline hazard function in Equation (6) such that *h*_03_(*t*_2_|*t*_1_) = *h*_03_(*t*_2_ − *t*_1_) [17]. The semi-Markov model assumes that the conditional baseline hazard for death at time *t*_2_ is dependent on the time from dementia to death given the onset of dementia at time *t*_1_, which is also called the ‘sojourn time’.

To characterize the relationship between the longitudinal and survival processes, the association structure, *W*_*g*_, can be formulated in various ways, which allows for a more tailored and flexible interpretation for the interplay between the two processes [20, 21]. We consider the following two commonly used association structures:

#### Current value (CV) association structure

The association structure *W*_*g*_, as defined in equations (4)-(6), has the following form:

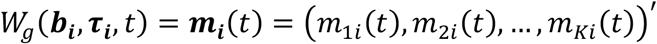

where *m*_*ki*_(*t*) denotes the vector of the true unobserved value of the *k*^th^ longitudinal measure at time *t* in Equations (1) and (2), *k* = 1,2,3, … *K*. Take the transition from the dementia state to the death state (Equation (6)) as an example, we assume the true values of all longitudinal measures at time *t*_2_, denoted as ***m***_***i***_(*t*_2_), are associated with the hazard for death at time *t*_2_ given that dementia occurred at time *t*_1_, denoted as *h*_3_(*t*_2_|*t*_1_).

#### Shared random effects (RE) association structure

As an alternative to the current value association structure, the association structure *W*_*g*_ in the shared random effects structure has the following form:

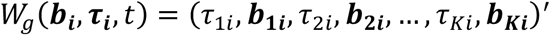

where *τ*_*ki*_ is the random changepoint and ***b***_***ki***_ is the vector of random intercepts and slopes for the *k*th longitudinal measure. This model posits that the hazard for dementia or death depends on the subject-specific changepoint and deviations from the average intercept and slope of the longitudinal sub-model.

If we assume two longitudinal measures (*K* = 2), the structure of our proposed joint model is displayed in **Fig. 3**. The random effects are crucial in constructing the joint model in the following three aspects. First, the correlation between the longitudinal measures is established through these random effects. Second, the longitudinal and survival processes are linked by the random effects directly in the shared random effects structure and indirectly in the current value structure. Third, the dependence structure between the non-terminal and terminal events are captured by the random effects.

**Fig. 3.**
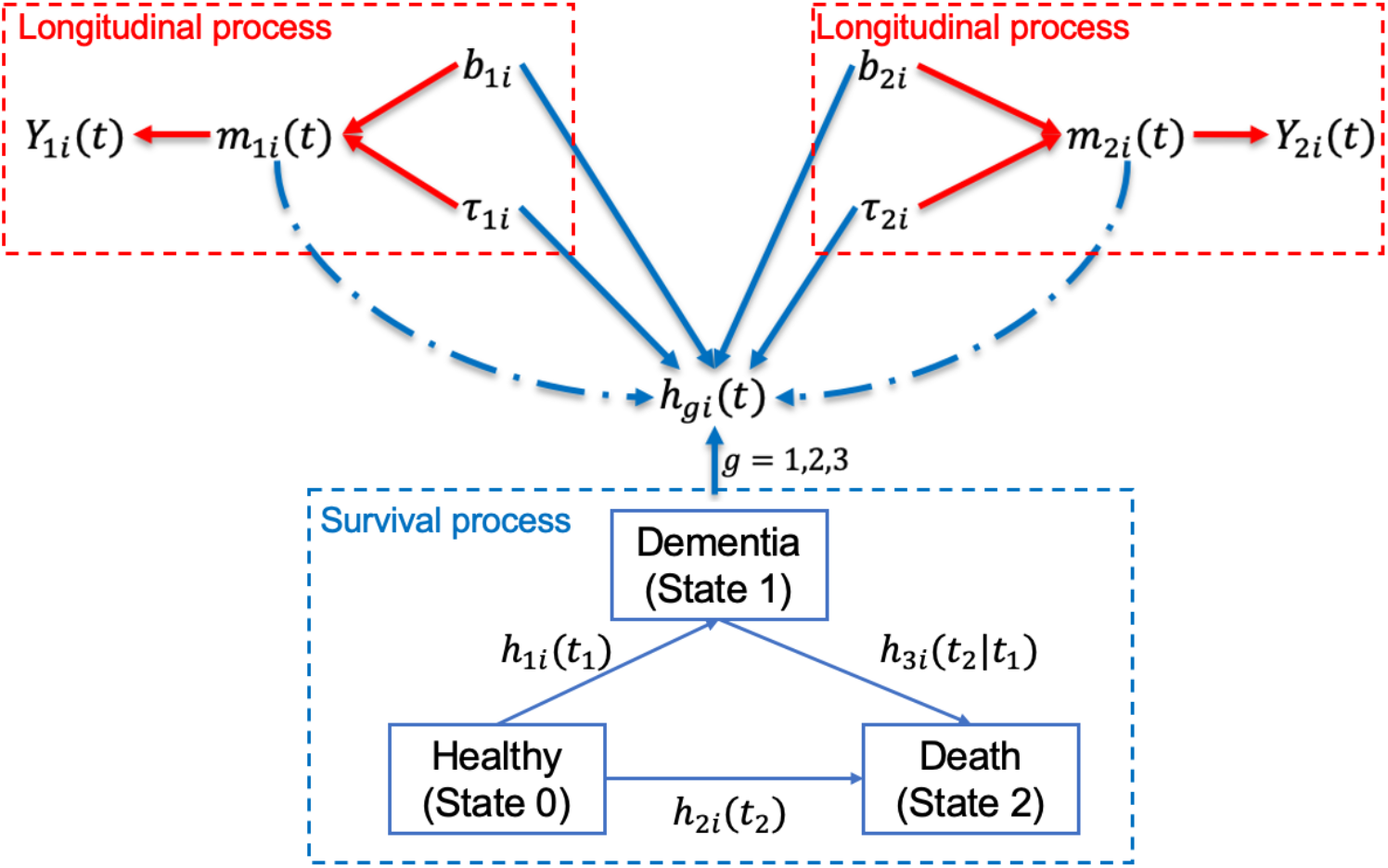
The proposed joint model framework. Two red boxes represent the longitudinal process for two longitudinal measures, and the blue box is the survival process. ***Y***_**1*i***_(***t***) and ***Y***_**2*i***_(***t***) are two longitudinal measures, ***m***_**1*i***_(***t***) and ***m***_**2*i***_(***t***) are the true unobserved values for two longitudinal measures, ***τ***_**1*i***_ and ***τ***_**2*i***_ are two correlated changepoints for ***Y***_**1*i***_(***t***) and ***Y***_**2*i***_(***t***), ***b***_**1*i***_ and ***b***_**2*i***_ are correlated random intercepts and slopes for ***Y***_**1*i***_(***t***) and ***Y***_**2*i***_(***t***). ***h***_***gi***_(***t***) denotes the hazard function for each health state transition in the survival process. Both longitudinal and survival processes are linked by either random effects (solid blue lines for the shared random effect model) or the true unobserved values of longitudinal measures (dashed blue lines for the current value model)

## 3 Model Estimation

### 3.1 Likelihood

The following data are observed for each participant *i*. Let ***Y***_***i***_ = (***Y***_**1*i***_, ***Y***_**2*i***_, …, ***Y***_***Ki***_) be the vector of *K* longitudinal measures. Let *C*_*i*_ be the right-censoring time for semi-competing risks. *T*_1*i*_ and *T*_2*i*_ are the times to the non-terminal (e.g., dementia) and terminal (e.g., death) events. *L*_1*i*_ = min {*C*_*i*_, *T*_1*i*_, *T*_2*i*_}, *L*_2*i*_ = min {*C*_*i*_, *T*_2*i*_}, Δ_1*i*_ = *I*{*T*_1*i*_ ≤ min(*T*_2*i*_, *C*_*i*_)} denotes the indicator of the non-terminal event, and Δ_2*i*_ = *I*{*T*_2*i*_ ≤ *C*_*i*_} denotes the indicator of the terminal event. Let ***D***_***i***_ = {*L*_1*i*_, Δ_1*i*_, *L*_2*i*_, Δ_2*i*_} be the set of observed time to event data. Let ***ψ*** be the full parameter vector. ***τ***_***i***_ and ***b***_***i***_ are vectors of random changepoints and random intercepts and slopes, respectively. We assume that the random effects underly both the longitudinal and survival processes, which means that the random effects account for the association between the longitudinal and survival outcomes (conditional independence) [22]. The likelihood is constructed under the commonly used assumption that both the longitudinal and survival processes are independent given the random effects: [20, 22, 23]

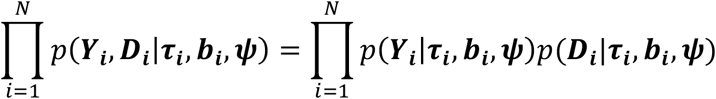

For the longitudinal part, we have

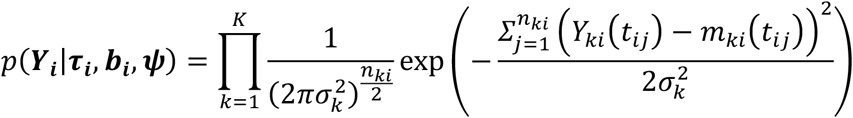

For the survival part, we construct the following likelihood

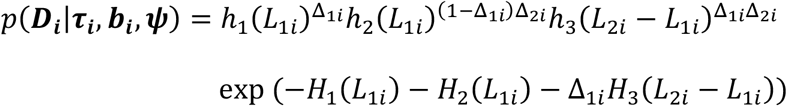

where *h*_*g*_(*t*) is the hazard function for each of the three transitions in **Fig. 2** and **Fig. 3**. 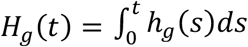 represents the cumulative hazard function. Note that we use Gauss-Legendre quadrature with 8 nodes to approximate the cumulative hazard function when the survival sub-model has a current value association structure. That is, 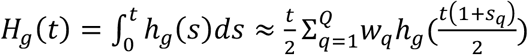, where *w*_*q*_ is the standardized weight for quadrature node *q* with *q* ∈ {1,2, …, *Q*}. *s*_*q*_ is the location for quadrature node *q*. The specific individual likelihood functions in the survival part for four specific cases (**Fig. 4**) are given as follows [19].

**Fig. 4.**
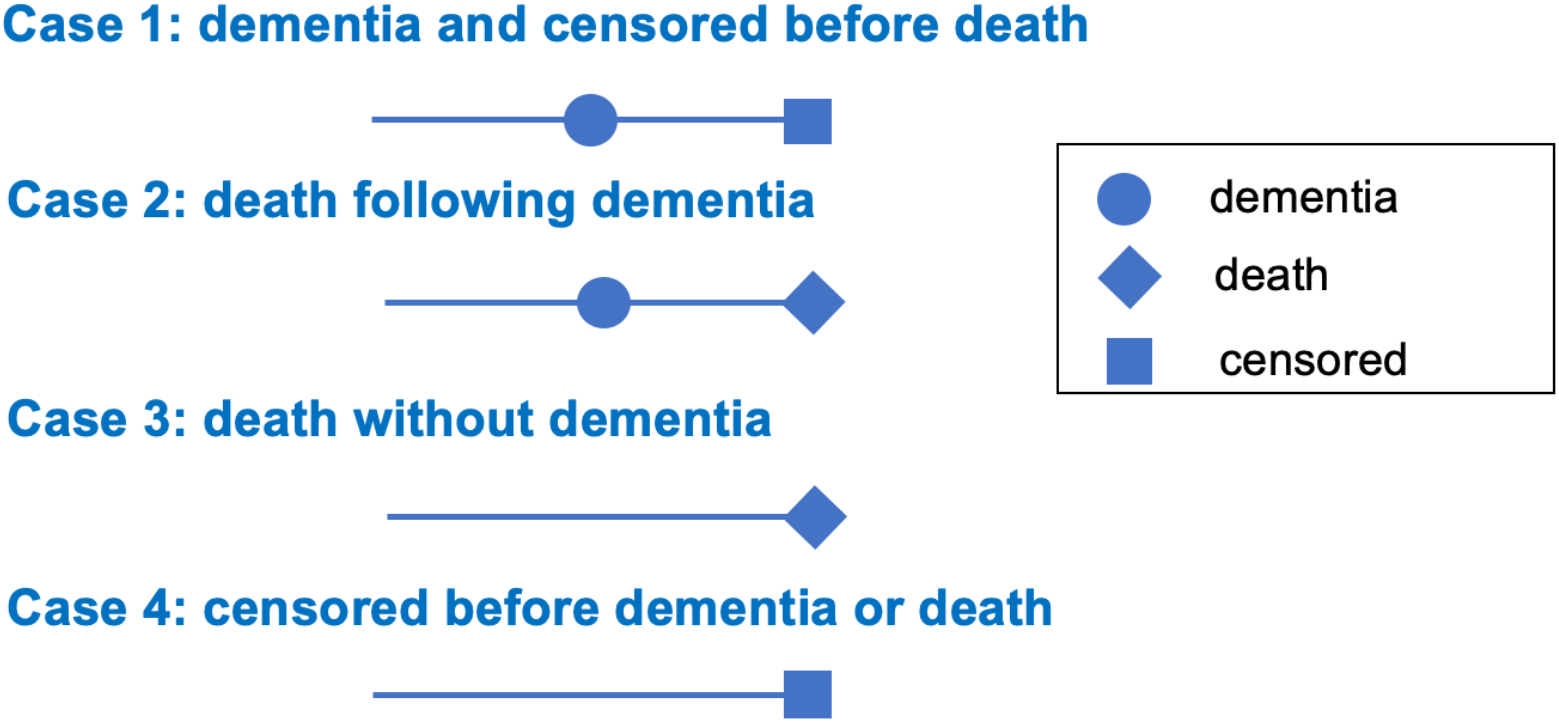
Four scenarios for dementia and death. The circle symbol represents the occurrence of dementia, the diamond symbol denotes the occurrence of death, and the square means the participant is censored

#### Case 1

Participant *i* develops dementia at time *L*_1*i*_ and is censored at time *C*_*i*_ before death:

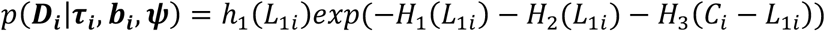

#### Case 2

Participant *i* is dead at time *L*_2*i*_, following the onset of dementia at time *L*_1*i*_:

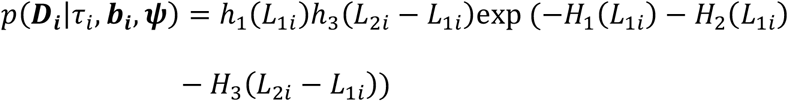

#### Case 3

Participant *i* is dead at time *L*_1*i*_ without developing dementia:

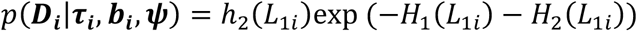

#### Case 4

Participant *i* is censored at time *C*_*i*_, prior to dementia or death:

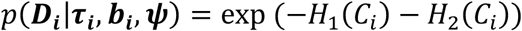

### 3.2 Bayesian Inference

We utilize a Bayesian approach for estimation since the joint model involves a large number of parameters with a complicated joint likelihood. The joint posterior distribution is analogous to:

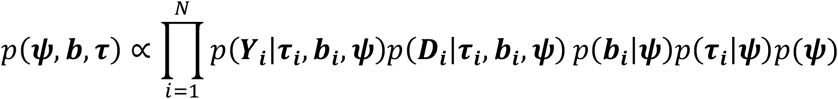

where *p*(***ψ***) is the joint prior distribution for the full parameter vector ***ψ***. We apply weakly informative priors to all the model parameters. The fixed regression coefficients, i.e., *β*_*k*0_, *β*_*k*1_, *β*_*k*2_, *α*_*g*_, and ***ζ***_***g***_ are assigned weakly informative normal priors with a large variance (i.e., 100). The fixed effects in the mean changepoints, i.e., *β*_*τk*0_, *β*_*τk*1_, and *γ*_*k*_, are assigned bounded uniform priors to prevent the estimated changepoint values from being biologically implausible. The parameter values in the uniform priors depend on the data. The residual standard deviations of the longitudinal measures *σ*_*k*_, are assigned a half Cauchy prior, i.e., *σ*_*k*_∼*half* − *Cauchy*(0, 2.5).

We decompose the random effects variance-covariance matrices using Cholesky decomposition [24]: Σ_*τ*_ = **Λ**_*τ*_Ω_*τ*_**Λ**_*τ*_ for random changepoints and Σ_*b*_ = **Λ**_*b*_Ω_b_**Λ**_*b*_ for random intercept and slopes, where **Λ**_*τ*_ and **Λ**_*b*_ denote the diagonal matrices containing standard deviation terms and Ω_*τ*_ and Ω_b_ are the correlation matrices. We employ the LKJ correlation priors parameterized in terms of its Cholesky factor with shape parameters 1 for Ω_*τ*_ and Ω_b_ and half-Cauchy priors (i.e., half-Cauchy (0, 2.5)) on the standard deviation terms in **Λ**_*τ*_ and **Λ**_*b*_.

The Bayesian estimation is implemented using the R interface of Stan with the R package rstan [25]. Stan utilizes a No-U-Turn Sampler version of Hamiltonian Monte Carlo (HMC) algorithm which optimizes the exploration of the target distribution based on Hamiltonian dynamics [26]. It has been shown that HMC has the capacity to handle complex nonlinear estimation tasks that might prove challenging for Gibbs sampling [27]. Additionally, the HMC algorithm implemented in Stan has proven to be effective in joint models with complex nonlinear parametrizations and random changepoint models [24, 27]. To assess model convergence, we use the potential scale reduction statistic 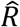 with 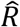 values < 1.1 for all parameters indicating successful model convergence. Widely Applicable Information Criterion (WAIC) is used to compare model fit [28]. WAIC is a useful tool for estimating pointwise out-of-sample prediction accuracy within the Bayesian framework. It is calculated based on the log-likelihood evaluated at the posterior simulations of the parameter values [29]. A smaller WAIC indicates a better model fit.

## 4 Simulation Study Design

In this section, we performed a simulation study to evaluate the estimation of changepoints and the associations between the longitudinal and survival processes in our proposed joint models with various changepoint formulations and association structures.

### 4.1 Data Generation

We simulated two correlated longitudinal measures (*K* = 2) and two semi-competing risks (e.g., dementia and death) for 1000 subjects in each simulated dataset. We chose the bent-cable model to simulate the two longitudinal measures since a smooth change aligns more closely with real-world scenarios in practice. Moreover, the bent-cable model is approximated to a piecewise model when the change zone is small enough. The two longitudinal measures *Y*_*ki*_(*t*_*ij*_), *k* = 1,2, were simulated from the following model:

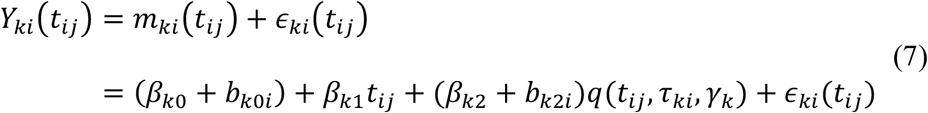

where *q*(*t*_*ij*_, *τ*_*ki*_, *γ*_*k*_) was described in Equation (3). The baseline time variable *t*_*i*0_ was generated from a uniform distribution *U*(−5, 5) to represent the baseline age subtracted by 60. The longitudinal measures were observed until the occurrence of either dementia or death or at the 20^th^ year of the follow-up period. We assumed that *Y*_1*i*_(*t*_*ij*_) had a narrow change zone (smaller *γ*_1_) around the changepoint, mimicking a longitudinal trajectory with a sharp change, while *Y*_2*i*_(*t*_*ij*_) exhibited a smooth change (larger *γ*_2_). The omission of the random slope before the changepoint was due to the flat and homogeneous trajectory pattern before the changepoint observed in real data. We further assumed that the correlation between the two longitudinal measures were fully captured by the random changepoints *τ*_*ki*_. The random intercepts and slopes across two longitudinal measures were generated with 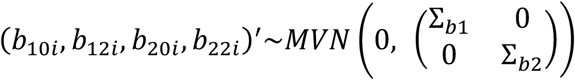 where 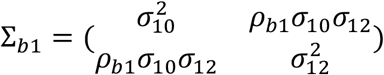 and 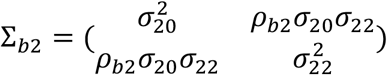. The random changepoints were 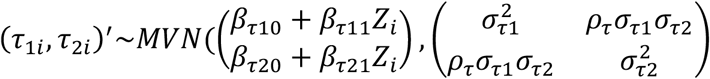 with *Z*_*i*_∼*Bernoulli*(0.2).

The times to semi-competing risks were generated under two scenarios of association structures, i.e., current value and shared random effects, to reflect the uncertainty about the true association structures in real-world scenarios. For the shared random effects structure, the semi-competing risks were generated based on the following survival sub-model:

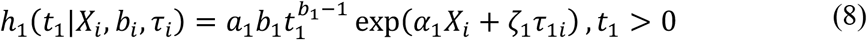

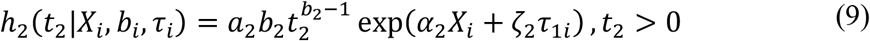

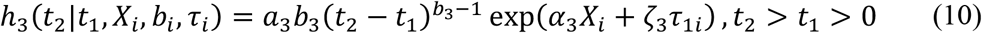

Random intercepts and slopes were excluded due to their insignificant effects on the survival part observed in the real data application. It has been suggested to include only one changepoint in the survival process if two longitudinal measures are highly correlated [13]. Because adding changepoints from two correlated longitudinal measures to the survival process can lead to multicollinearity, which weakens statistical power of the joint model. Besides, including too many variables or random effects is impractical since it slows the estimation process of an already complex joint model and can lead to convergence issues. Therefore, only the changepoint in the first longitudinal measure, *τ*_1*i*_, was included in the survival sub-model to avoid multicollinearity since the two changepoints were assumed to be highly correlated based on our real data application. Similarly, for the current value association structure, the semi-competing risks were derived using the following survival sub-model

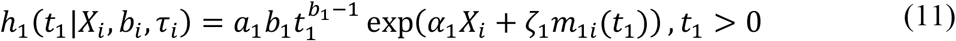

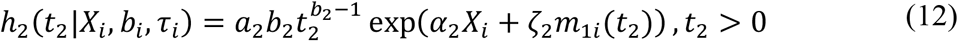

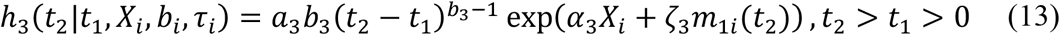

We included the true unobserved value for the first longitudinal measure at the same time of the event, denoted as *m*_1*i*_(*t*_1_) or *m*_1*i*_(*t*_2_), in the association structure to avoid multicollinearity. In both scenarios, *X*_*i*_ was generated from Bernoulli(0.5), and the semi-competing risks data were simulated based on the cause-specific hazard approach described in Beyersmann et al. (2009) and Bender et al. (2005) [30, 31]. Detailed steps for simulating times to semi-competing risks were described in **Supplementary Materials A**.

### 4.2 Simulation Scenarios

For each of the association structures from the previous section, we considered two cohort types: a disease cohort which has an earlier changepoint occurrence, a higher dementia rate, and more frequent visits, and a community cohort which has a later changepoint occurrence, a lower dementia rate, and less frequent visits. Therefore, we simulated four scenarios in total:

- **Scenario 1**: disease cohort with shared random effects (RE) association structure.
- **Scenario 2**: disease cohort with current value (CV) association structure.
- **Scenario 3**: community cohort with RE association structure.
- **Scenario 4**: community cohort with CV association structure.

The parameters used to generate our simulated datasets were based on estimates from the Framingham Heart Study (FHS) Offspring cohort data described in Section 6 to allow for reasonable generalization. Note that in the community cohort (**Scenarios 3** and **4**), only a portion of the participants experienced the observed changepoints. Main parameter values used to simulate data, event rates, and time interval between two adjacent longitudinal visits were displayed in **Table 1**. For each scenario, 200 datasets were generated and analyzed.

**Table 1.** Main parameter settings for all four scenarios in the simulation study.

| Parameters | Scenario 1 | Scenario 2 | Scenario 3 | Scenario 4 |
| --- | --- | --- | --- | --- |
| **Longitudinal** |  |  |  |  |
| Interval between visits | 2 years | 2 years | 5 years | 5 years |
| $\beta_{10}$ | -0.18 | -0.18 | -0.18 | -0.18 |
| $\beta_{11}$ | -0.02 | -0.02 | -0.02 | -0.02 |
| $\beta_{12}$ | -0.15 | -0.15 | -0.15 | -0.15 |
| $\gamma_1$ | 2 | 2 | 2 | 2 |
| $\sigma_{10}$ | 0.4 | 0.4 | 0.4 | 0.4 |
| $\sigma_{12}$ | 0.05 | 0.05 | 0.05 | 0.05 |
| $\rho_{b1}$ | 0.3 | 0.3 | 0.3 | 0.3 |
| $\beta_{20}$ | -0.03 | -0.03 | -0.03 | -0.03 |
| $\beta_{21}$ | -0.02 | -0.02 | -0.02 | -0.02 |
| $\beta_{22}$ | -0.2 | -0.2 | -0.2 | -0.2 |
| $\gamma_2$ | 4 | 4 | 4 | 4 |
| $\sigma_{20}$ | 0.5 | 0.5 | 0.5 | 0.5 |
| $\sigma_{22}$ | 0.05 | 0.05 | 0.05 | 0.05 |
| $\rho_{b2}$ | 0.3 | 0.3 | 0.3 | 0.3 |
| **Changepoint** |  |  |  |  |
| $\beta_{\tau 10}$ | 15 | 15 | 25 | 25 |
| $\beta_{\tau 11}$ | -3 | -3 | -3 | -3 |
| $\sigma_{\tau 1}$ | 3 | 3 | 8 | 8 |
| $\beta_{\tau 20}$ | 12 | 12 | 27 | 27 |
| $\beta_{\tau 21}$ | -5 | -5 | -5 | -5 |
| $\sigma_{\tau 2}$ | 3 | 3 | 8 | 8 |
| $\rho_{\tau}$ | 0.8 | 0.8 | 0.8 | 0.8 |
| **Survival** |  |  |  |  |
| Rates for dementia, death without dementia, and death after dementia | 30%; 15%; 20% | 30%; 15%; 20% | 10%; 10%; 6% | 15%; 10%; 10% |
| $\alpha_1$ | -0.6 | -0.2 | -0.6 | -0.2 |
| $\zeta_1$ | -0.3 | -2.5 | -0.3 | -2.5 |
| $\alpha_2$ | -0.6 | -0.4 | -0.6 | -0.4 |
| $\zeta_2$ | -0.1 | -0.6 | -0.1 | -0.6 |
| $\alpha_3$ | -0.1 | -0.2 | -0.1 | -0.2 |
| $\zeta_3$ | -0.1 | -0.5 | -0.1 | -0.5 |

Scenario 1: disease cohort with shared random effects association structure; Scenario 2: disease cohort with current value association structure; Scenario 3: community cohort with shared random effects association structure; Scenario 4: community cohort with current value association structure

### 4.3 Analysis Models

The selection of an appropriate model is based not only on model criterion but also on underlying rationales. The piecewise model may be preferred due to its straightforward interpretation and lower computational burden compared to the bent-cable model. The current value structure could be selected if the focus is on understanding the survival process, while the shared random effect structure is chosen when the focus is correcting longitudinal bias [20].

Therefore, to analyze each of the four scenarios described in **Section 4.2**, we evaluated four joint models with different combinations of changepoint formulations and association structures described in the previous sections:

- **Model BC+RE**: bent-cable model (Equation (7)) + shared random effects association structure (Equations (8)-(10)).
- **Model PW+RE**: piecewise model + shared random effects association structure (Equations (8)-(10)). Similar to Equation (7), the piecewise model in the longitudinal part is specified in the following equation by replacing the smooth function *q*(*t*_*ij*_, *τ*_*ki*_, *γ*_*k*_) with identity function *I*(*t*_*ij*_ − *τ*_*ki*_):

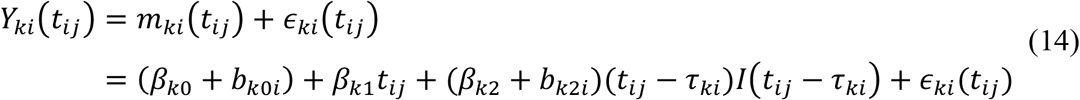
- **Model BC+CV**: bent-cable model (Equation (7)) + current value association structure (Equations (11)-(13)).
- **Model PW+CV**: piecewise model (Equation (14)) + current value association structure (Equations (11)-(13)).

**Fig. S1** provided an illustration of the simulation scenarios and analysis models.

Weakly informative priors were specified in **Section 3.2**. The reasonable ranges for the changepoint parameters were specified by the following uniform priors: mean changepoint *β*_*τ*10_ and *β*_*τ*20_ ∼ *U*(10,20) for the disease cohort scenarios and *U*(20,30) for the community cohort scenarios; the effect of covariate *Z*_*i*_ on mean changepoints *β*_*τ*11_ and *β*_*τ*21_∼*U*(−10,0); the half width of the change zone *γ*_1_ and *γ*_2_∼*U*(0,6). We obtained 6000 MCMC sample iterations from each of the 3 parallel chains with the first 3000 iterations as a warm-up phase. Convergence was evaluated through the potential scale reduction statistic. We calculated and compared percent bias and 95% coverage probabilities for each parameter based on posterior mean, along with WAIC values for each model.

## 5 Simulation Results

Our primary interests were the estimated mean changepoints (*β*_*τ*10_, *β*_*τ*20_) and slope difference before and after the changepoint (*β*_12_ and *β*_22_) in the longitudinal sub-model, as well as the association parameters (*ζ*_1_, *ζ*_2_, *ζ*_3_) in the survival sub-model, especially the impact of the longitudinal process on the hazard of death conditional on dementia onset (*ζ*_3_). The posterior means for *β*_*τ*10_, *β*_*τ*20_, and *ζ*_3_ in all four scenarios were illustrated in **Fig. 5** and **Fig. 6**. Numerical results were provided in **Table 2** for **Scenario 1** and **Tables S1-3** for **Scenarios 2-4**.

**Table 2.** Simulation results for Scenario 1 (disease cohort with shared random effects association structure).

| Parameter | True | BC+RE |  |  | PW+RE |  |  | BC+CV* |  |  | PW+CV* |  |  |
| --- | --- | --- | --- | --- | --- | --- | --- | --- | --- | --- | --- | --- | --- |
|  |  | Mean | PB | CP | Mean | PB | CP | Mean | PB | CP | Mean | PB | CP |
| Longitudinal |  |  |  |  |  |  |  |  |  |  |  |  |  |
| $\beta_{10}$ | -0.18 | -0.18 | -0.24 | 0.95 | -0.18 | -0.99 | 0.95 | -0.18 | -0.39 | 0.96 | -0.18 | -1.38 | 0.96 |
| $\beta_{11}$ | -0.02 | -0.02 | 0.02 | 0.94 | -0.02 | 3.02 | 0.89 | -0.02 | 0.29 | 0.95 | -0.02 | 4.08 | 0.81 |
| $\beta_{12}$ | -0.15 | -0.15 | 1.81 | 0.94 | -0.14 | -4.56 | 0.72 | -0.16 | 6.62 | 0.79 | -0.15 | -1.49 | 0.89 |
| $\beta_{20}$ | -0.03 | -0.03 | 4.67 | 0.91 | -0.03 | -2.73 | 0.93 | -0.03 | 3.23 | 0.90 | -0.03 | -3.12 | 0.92 |
| $\beta_{21}$ | -0.02 | -0.02 | -0.42 | 0.95 | -0.02 | 22.45 | 0.00 | -0.02 | -0.09 | 0.95 | -0.02 | 23.36 | 0.00 |
| $\beta_{22}$ | -0.20 | -0.20 | 0.43 | 0.96 | -0.18 | -11.65 | 0.00 | -0.20 | 1.15 | 0.92 | -0.18 | -11.32 | 0.00 |
| $\gamma_1$ | 2.00 | 2.03 | 1.58 | 0.94 | - | - | - | 2.43 | 21.40 | 0.87 | - | - | - |
| $\gamma_2$ | 4.00 | 4.04 | 0.99 | 0.96 | - | - | - | 4.12 | 3.03 | 0.93 | - | - | - |
| Changepoint |  |  |  |  |  |  |  |  |  |  |  |  |  |
| $\beta_{\tau 10}$ | 15.00 | 15.09 | 0.60 | 0.92 | 14.80 | -1.33 | 0.82 | 15.47 | 3.13 | 0.63 | 15.11 | 0.75 | 0.91 |
| $\beta_{\tau 20}$ | 12.00 | 12.02 | 0.21 | 0.97 | 11.55 | -3.78 | 0.08 | 12.13 | 1.06 | 0.85 | 11.64 | -3.03 | 0.26 |
| $\beta_{\tau 11}$ | -3.00 | -2.99 | -0.44 | 0.95 | -2.98 | -0.80 | 0.95 | -3.07 | 2.34 | 0.91 | -3.08 | 2.63 | 0.92 |
| $\beta_{\tau 21}$ | -5.00 | -5.01 | 0.18 | 0.94 | -4.87 | -2.64 | 0.90 | -5.05 | 0.95 | 0.93 | -4.91 | -1.80 | 0.93 |
| $\sigma_{\tau 1}$ | 3.00 | 3.03 | 1.07 | 0.96 | 3.02 | 0.59 | 0.97 | 3.11 | 3.63 | 0.90 | 3.10 | 3.43 | 0.91 |
| $\sigma_{\tau 2}$ | 3.00 | 3.00 | 0.12 | 0.93 | 2.97 | -1.01 | 0.93 | 3.03 | 1.01 | 0.93 | 3.00 | -0.10 | 0.92 |
| $\rho_{\tau}$ | 0.80 | 0.80 | -0.62 | 0.97 | 0.80 | -0.35 | 0.97 | 0.79 | -0.64 | 0.97 | 0.79 | -1.27 | 0.98 |
| Survival |  |  |  |  |  |  |  |  |  |  |  |  |  |
| $\alpha_1$ | -0.60 | -0.59 | -1.50 | 0.94 | -0.59 | -1.62 | 0.95 | -0.52 | - | - | -0.51 | - | - |
| $\zeta_1$ | -0.30 | -0.30 | 0.12 | 0.95 | -0.31 | 3.79 | 0.96 | -0.68 | - | - | -0.68 | - | - |
| $\alpha_2$ | -0.60 | -0.58 | -2.92 | 0.93 | -0.58 | -2.70 | 0.93 | -0.56 | - | - | -0.56 | - | - |
| $\zeta_2$ | -0.10 | -0.10 | 0.12 | 0.95 | -0.11 | 9.96 | 0.93 | -0.20 | - | - | -0.21 | - | - |
| $\alpha_3$ | -0.10 | -0.10 | 1.48 | 0.93 | -0.10 | -1.94 | 0.93 | -0.08 | - | - | -0.08 | - | - |
| $\zeta_3$ | -0.10 | -0.11 | 7.26 | 0.95 | -0.11 | 7.48 | 0.93 | -0.14 | - | - | -0.14 | - | - |

True: parameter true value; Mean: posterior mean; PB: percent bias; CP: coverage probability; BC: bent-cable model; RE: shared random effects association structure; PW: piecewise model; CV: current value association structure.
\*Percent bias and coverage probability are not available for association parameters in the survival sub-model for Model BC+CV and PW+CV since the true association structure is RE.

**Fig. 5.**
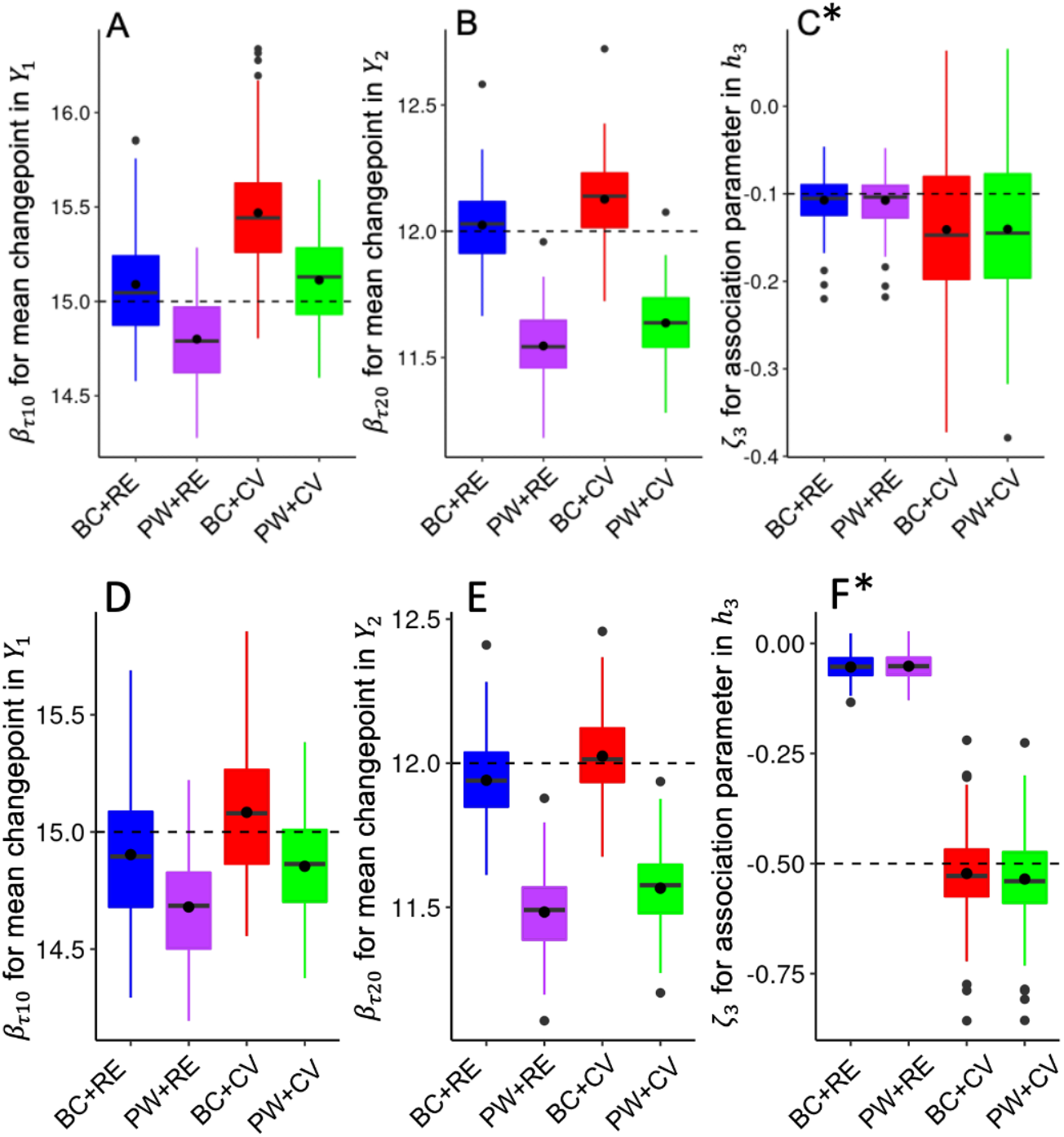
Distributions of posterior means for mean changepoints and association parameters in the disease cohort (Scenario 1 disease cohort with shared random effects structure and Scenario 2 disease cohort with current value structure). The upper three panels A, B, and C represent Scenario 1, while the bottom panels D, E, and F indicate Scenario 2. In each panel, the boxes denote the posterior means across all 200 simulated datasets based on each method, while the dashed line indicates the true value for the parameter. Panels A&D: posterior means for ***β***_***τ*10**_ (the mean changepoint parameter in ***Y***_**1**_ with an abrupt change); Panels B&E: posterior means for ***β***_***τ*20**_ (the mean changepoint parameter in ***Y***_**2**_ with a smooth change); Panels C&F: posterior means for ***ζ***_**3**_ (the association parameter between longitudinal and survival processes in ***h***_**3**_). Scenario 1: disease cohort with shared random effects association structure; Scenario 2: disease cohort with current value association structure; PW: piecewise model; BC: bent-cable model; RE: shared random effects association structure; CV: current value association structure *In Panels C&F, the results for ***ζ***_**3**_ from RE and CV models are not directly comparable, since ***ζ***_**3**_ has different interpretations in models with these two association structures. We mainly focus on the directionality of ***ζ***_**3**_.

**Fig. 6.**
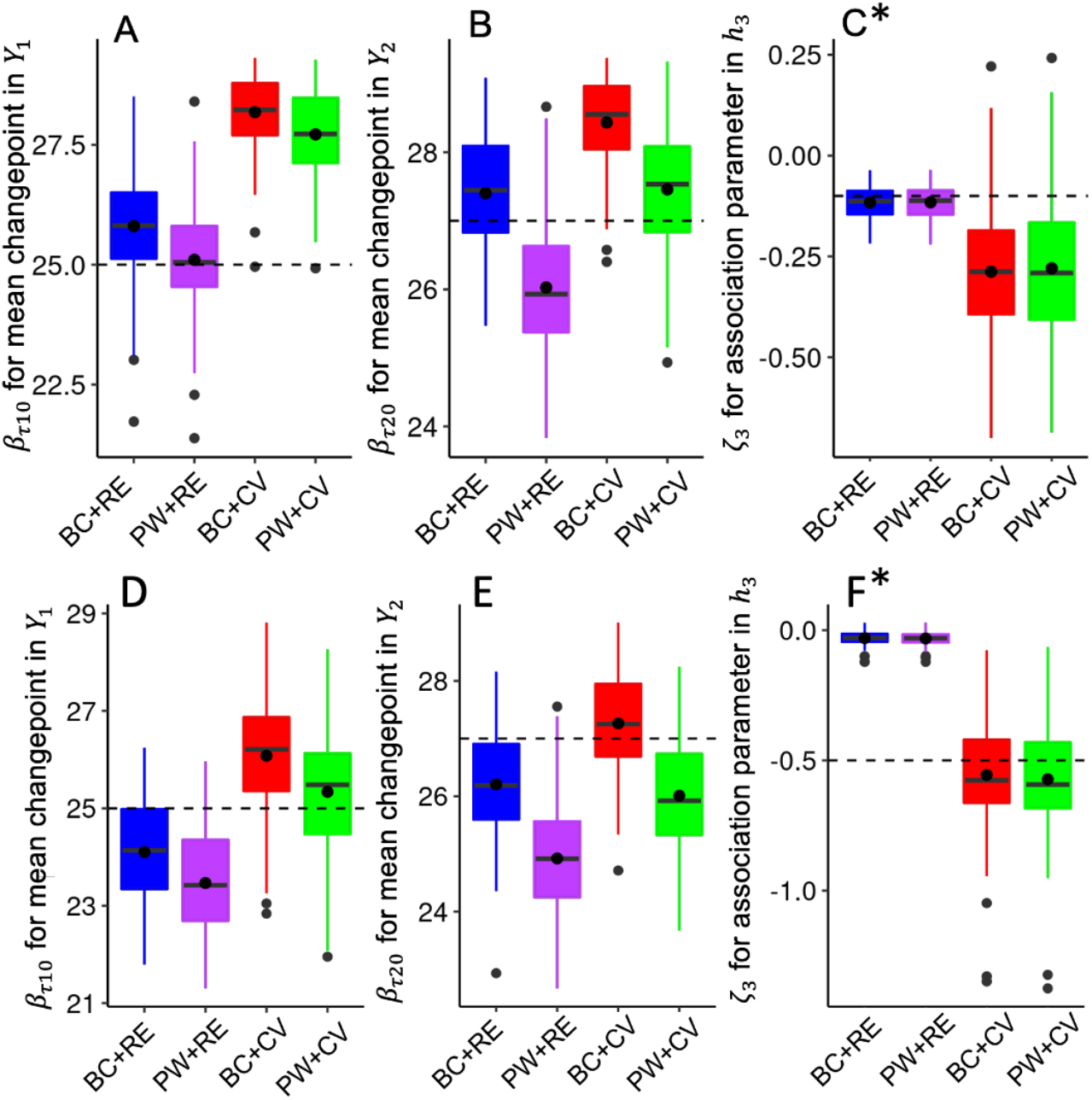
Distributions of posterior means for mean changepoints and association parameters in the community cohort (Scenario 3 community cohort with shared random effects structure and Scenario 4 community cohort with current value structure). The upper three panels A, B, and C represent Scenario 3, while the bottom panels D, E, and F indicate Scenario 4. In each panel, the boxes denote the posterior means across all 200 simulated datasets based on each method, while the dashed line indicates the true value for the parameter. Panels A&D: posterior means for ***β***_***τ*10**_ (the mean changepoint parameter in ***Y***_**1**_ with an abrupt change); Panels B&E: posterior means for ***β***_***τ*20**_ (the mean changepoint parameter in ***Y***_**2**_ with a smooth change); Panels C&F: posterior means for ***ζ***_**3**_ (the association parameter between longitudinal and survival processes in ***h***_**3**_). Scenario 3: community cohort with shared random effects association structure; Scenario 4: community cohort with current value association structure; PW: piecewise model; BC: bent-cable model; RE: shared random effects association structure; CV: current value association structure. *In Panels C&F, the results for *ζ*_3_ from RE and CV models are not directly comparable, since *ζ*_3_ has different interpretations in models with these two association structures. We mainly focus on the directionality of *ζ*_3_.

For **Scenario 1** RE structure in a disease cohort with relatively frequent longitudinal visits and higher event rates (**Panels A, B**, and **C** in **Fig. 5, Table 2**), the joint models that included the bent-cable model, which allowed for a smooth change, along with the correctly specified association structure (**Model BC+RE**), showed the best performance regarding bias (<5% for most parameters) and coverage probability (around 95%). These findings demonstrated the flexibility of the bent-cable model in handling changepoints in longitudinal measures that exhibited either an abrupt or smooth change with sufficient data. Parameter estimates from other models deviated from the true values, but the bias and coverage probability values for most parameters were within an acceptable range. When evaluating the impact of replacing the bent-cable model with a more parsimonious piecewise model (**Model BC+RE** vs **PW+RE** and **Model BC+CV** vs **PW+CV**), the piecewise model assuming an abrupt change produced a slower cognitive decline after the changepoint and earlier detection of the changepoint compared to the bent-cable model. Differences in the estimates were more pronounced for the cognitive measure with a smooth change *Y*_2_. The estimated survival parameters (*ζ*_1_, *ζ*_2_, and *ζ*_3_) demonstrated robustness, maintaining stability across two longitudinal specifications. When comparing two association structures in the survival sub-model (**Model BC+RE** vs **BC+CV** and **Model PW+RE** vs **PW+CV**), the current value structure resulted in a more drastic cognitive decline after the changepoint and a later occurrence of the changepoint compared to the shared random effects structure. Since the association parameters in two association structures had different model parametrizations and interpretations so they were not directly comparable. However, the same association directionality between the longitudinal and survival processes were captured even when the association structure was misspecified (**Panel C** in **Fig. 5**).

In **Scenario 2** disease cohort with CV structure (**Panels D, E, and F in Fig. 5, Table S1**), the joint model with a smooth change and the correctly specified association structure was **Model BC+CV**, which yielded the best performance. All other conclusions drawn from **Scenario 1** were also applicable. According to **Table S4**, the WAIC values for the first two scenarios showed that the correctly specified joint model with a smooth change (**Model BC+RE** in **Scenario 1** and **Model BC+CV** in **Scenario 2**) produced the smallest WAIC, indicating the best predictive accuracy for model fit.

For **Scenario 3** RE structure in a community cohort with relatively wide visit intervals and lower event rates (**Panels A, B**, and **C in Fig. 6, Table S2**), more bias was observed compared to the disease cohort overall. The joint models that included the bent-cable model and the correctly specified association structure (**Model BC+RE**) produced the best performance for the cognitive measure with a smooth change (*Y*_2_). In contrast, the piecewise model with the correctly specified association structure (**Model PW+RE**) showed optimal results for the cognitive measure with a sharp change (*Y*_1_). Additionally, both piecewise and bent-cable models showed comparable WAIC. These results indicated that the bent-cable model failed to improve upon the more parsimonious piecewise model in the community cohort. **Scenario 4** community cohort with CV structure **(Panels D, E**, and **F in Fig. 6, Table S3)** yielded similar conclusions to **Scenario 3**. Notably, in **Scenario 4**, the correctly specified association structure was the current value structure in **Model BC+CV** and **Model PW+CV**. Most of the findings in the disease cohort regarding model comparison applied to the community cohort, but the differences among models became more obvious in the community cohort.

## 6 Real Data Application

In this section, we used longitudinal cognitive data along with semi-competing risks data for dementia and death in a community cohort to illustrate our proposed models.

### 6.1 Data Description

The Framingham Heart Study (FHS) is a multigenerational cohort study initiated in 1948 by enrolling 5209 residents in the Original cohort [32]. In 1971, 5214 participants who were the offspring of the Original cohort and the spouses of these offspring were included in the Offspring cohort [33]. Beginning in 1999, the surviving participants in the FHS Offspring cohort were invited to join an ancillary study, where they were administered a battery of neuropsychological (NP) tests every four years [2]. Participants identified as having potential cognitive impairment were invited to undergo additional, annual neurologic and NP tests. NP tests were used to measure participants’ cognitive changes over time in four domains including memory and language [34]. A complete list of tests included in the memory and language domains were summarized in **Table S5**. For each participant at each visit, we computed a memory z-score and a language z-score by averaging the z-scores from all individual NP tests within the memory domain and the language domain, respectively. A dementia review panel with at least one neurologist and one neuropsychologist determined whether the participants had dementia, as well as the dementia type and the date of onset [2, 35]. We excluded the participants who had dementia onset before the age of 60 at baseline and had less than three visits with NP tests in both memory and language domains. Our final sample included 1117 participants with 4162 observations. Of these 1117 participants, 20 were diagnosed with dementia then censored during follow-up, 66 died following the diagnosis of dementia, 165 died without the onset of dementia, and 866 were censored without any events. The protocol of the FHS was approved by the Institutional Review Board at the Boston University Medical Campus.

### 6.2 Model Specification

We utilized z-scores for NP tests in the memory and language domains as two longitudinal measures, with dementia and death considered as semi-competing risks. Like the simulation study, we considered four models with two changepoint formulations and two association structures. Specifically, the longitudinal sub-models with two changepoint formulations were:

#### Piecewise model

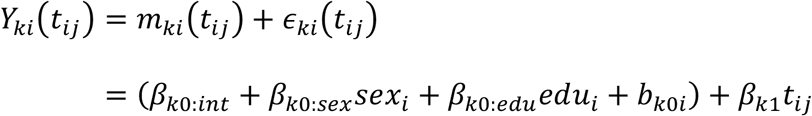

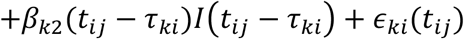

#### Bent-cable model

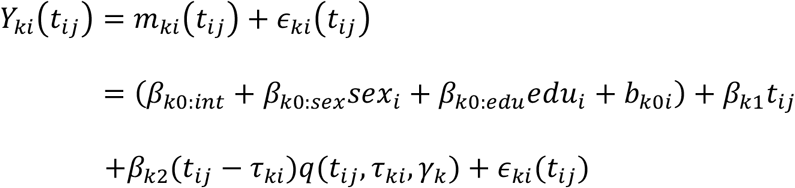

where *t*_*ij*_ denoted the age centered at 60 years for participant *i* at visit *j. Y*_*ki*_(*t*_*ij*_) represented memory z-scores for *k* = 1, and language z-scores for *k* = 2. We further included sex (*sex*_*i*_) and education years (*edu*_*i*_) as covariates. To reduce model complexity and facilitate convergence, we only included random effects in the intercept term (*b*_*k*0*i*_) and the changepoint term (*τ*_*ki*_), such that 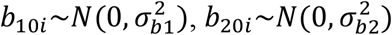, and 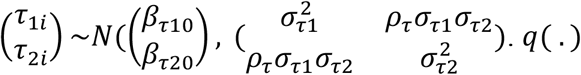 was the function for a smooth change around the changepoint defined in Equation (3).

For the survival sub-model, we considered two association structures in the following illness-death model:

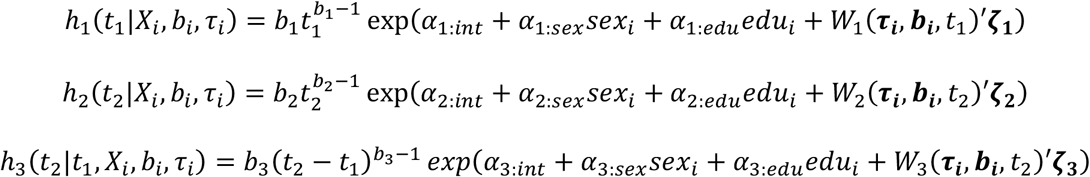

where *W*_*g*_(***τ***_***i***_, ***b***_***i***_, *t*)^′^***ζ***_***g***_ = *ζ*_*g*1_(*τ*_1*i*_ − 20) + *ζ*_*g*2_*b*_10*i*_ for the shared random effects structure and *W*_*g*_(***τ***_***i***_, ***b***_***i***_, *t*)^′^***ζ***_***g***_ = *ζ*_*g*1_*m*_1*i*_(*t*) for the current value structure, *g* ∈ {1,2,3}. Note that compared to the simulation study, we included sex and education years as covariates and incorporated both changepoints and random intercept in the shared random effects structure. The random changepoint was centered by subtracting 20 for better model convergence.

### 6.3 Results

Numerical results, including posterior means and 95% credible intervals (CI), were displayed in **Table 3**. Based on the WAIC values of these four models, **Model BC+RE** resulted in the best model fit, which suggested a shared random effects data structure for the FHS data. Therefore, for brevity, we mainly referred to **Model BC+RE** for the following parameter interpretations.

**Table 3.** Joint model results for the FHS Offspring cohort data.

| Param | BC+RE<br>(WAIC=6136) |  |  | PW+RE<br>(WAIC=6193) |  |  | BC+CV<br>(WAIC=6214) |  |  | PW+CV<br>(WAIC=6283) |  |  |
| --- | --- | --- | --- | --- | --- | --- | --- | --- | --- | --- | --- | --- |
|  | Mean | Lower | Upper | Mean | Lower | Upper | Mean | Lower | Upper | Mean | Lower | Upper |
| **Longitudinal** |  |  |  |  |  |  |  |  |  |  |  |  |
| $\beta_{10:int}$ | -0.18 | -0.27 | -0.08 | -0.18 | -0.27 | -0.08 | -0.18 | -0.28 | -0.08 | -0.19 | -0.28 | -0.09 |
| $\beta_{10:sex}$ | 0.28 | 0.22 | 0.34 | 0.28 | 0.22 | 0.33 | 0.28 | 0.22 | 0.34 | 0.28 | 0.22 | 0.34 |
| $\beta_{10:edu}$ | 0.08 | 0.07 | 0.10 | 0.08 | 0.07 | 0.10 | 0.08 | 0.07 | 0.10 | 0.08 | 0.07 | 0.10 |
| $\beta_{11}$ | -0.01 | -0.01 | -0.01 | -0.01 | -0.01 | -0.01 | -0.01 | -0.01 | -0.01 | -0.01 | -0.01 | -0.01 |
| $\beta_{12}$ | -0.17 | -0.20 | -0.15 | -0.15 | -0.17 | -0.14 | -0.17 | -0.20 | -0.15 | -0.16 | -0.18 | -0.14 |
| $\beta_{20:int}$ | -0.04 | -0.14 | 0.06 | -0.03 | -0.13 | 0.06 | -0.04 | -0.14 | 0.07 | -0.03 | -0.14 | 0.06 |
| $\beta_{20:sex}$ | 0.10 | 0.04 | 0.16 | 0.10 | 0.04 | 0.16 | 0.10 | 0.04 | 0.16 | 0.10 | 0.04 | 0.16 |
| $\beta_{20:edu}$ | 0.14 | 0.12 | 0.15 | 0.14 | 0.12 | 0.15 | 0.14 | 0.12 | 0.15 | 0.14 | 0.12 | 0.15 |
| $\beta_{21}$ | -0.01 | -0.01 | 0.00 | -0.01 | -0.01 | 0.00 | -0.01 | -0.01 | 0.00 | -0.01 | -0.01 | 0.00 |
| $\beta_{22}$ | -0.19 | -0.24 | -0.15 | -0.12 | -0.15 | -0.10 | -0.19 | -0.25 | -0.14 | -0.13 | -0.15 | -0.10 |
| $\gamma_1$ | 6.18 | 3.21 | 7.91 | - | - | - | 4.94 | 2.28 | 7.57 | - | - | - |
| $\gamma_2$ | 7.44 | 6.17 | 7.98 | - | - | - | 7.39 | 5.98 | 7.98 | - | - | - |
| $\sigma_{b1}$ | 0.44 | 0.41 | 0.46 | 0.44 | 0.41 | 0.46 | 0.44 | 0.42 | 0.46 | 0.44 | 0.42 | 0.46 |
| $\sigma_{b2}$ | 0.49 | 0.46 | 0.51 | 0.48 | 0.46 | 0.51 | 0.49 | 0.47 | 0.51 | 0.48 | 0.46 | 0.51 |
| **Changepoint** |  |  |  |  |  |  |  |  |  |  |  |  |
| $\beta_{\tau 10}$ | 25.82 | 23.99 | 27.96 | 25.43 | 23.61 | 27.59 | 27.64 | 25.40 | 30.17 | 27.51 | 25.21 | 30.05 |
| $\beta_{\tau 20}$ | 30.00 | 27.80 | 32.34 | 27.90 | 25.70 | 30.36 | 31.39 | 28.91 | 34.11 | 29.38 | 26.94 | 32.21 |
| $\sigma_{\tau 1}$ | 9.54 | 8.45 | 10.86 | 9.72 | 8.52 | 11.18 | 10.46 | 9.16 | 11.98 | 10.65 | 9.31 | 12.23 |
| $\sigma_{\tau 2}$ | 8.97 | 7.74 | 10.45 | 9.63 | 8.19 | 11.35 | 9.54 | 8.17 | 11.25 | 10.20 | 8.61 | 12.24 |
| $\rho_{\tau}$ | 0.90 | 0.85 | 0.94 | 0.90 | 0.84 | 0.94 | 0.90 | 0.85 | 0.95 | 0.90 | 0.84 | 0.95 |
| **Survival** |  |  |  |  |  |  |  |  |  |  |  |  |
| $\alpha_{1:sex}$ | -0.60 | -1.16 | -0.07 | -0.60 | -1.15 | -0.06 | -0.01 | -0.53 | 0.49 | -0.01 | -0.55 | 0.54 |
| $\alpha_{1:edu}$ | -0.14 | -0.25 | -0.04 | -0.14 | -0.25 | -0.04 | 0.04 | -0.06 | 0.14 | 0.04 | -0.06 | 0.14 |
| $\zeta_1$ | -0.30 | -0.37 | -0.23 | -0.29 | -0.36 | -0.23 | -2.53 | -3.10 | -2.02 | -2.55 | -3.17 | -2.01 |
| $\alpha_{2:sex}$ | -0.59 | -0.92 | -0.26 | -0.58 | -0.91 | -0.25 | -0.42 | -0.73 | -0.12 | -0.43 | -0.75 | -0.12 |
| $\alpha_{2:edu}$ | -0.01 | -0.07 | 0.05 | -0.01 | -0.08 | 0.05 | 0.04 | -0.02 | 0.10 | 0.04 | -0.02 | 0.10 |
| $\zeta_2$ | -0.08 | -0.12 | -0.04 | -0.07 | -0.12 | -0.03 | -0.62 | -0.91 | -0.32 | -0.60 | -0.89 | -0.30 |
| $\alpha_{3:sex}$ | -0.08 | -0.63 | 0.46 | -0.08 | -0.62 | 0.46 | -0.21 | -0.73 | 0.33 | -0.20 | -0.72 | 0.33 |
| $\alpha_{3:edu}$ | 0.12 | 0.01 | 0.23 | 0.12 | 0.01 | 0.24 | 0.07 | -0.04 | 0.17 | 0.06 | -0.04 | 0.17 |
| $\zeta_3$ | 0.06 | 0.02 | 0.10 | 0.06 | 0.02 | 0.10 | 0.29 | -0.11 | 0.69 | 0.31 | -0.12 | 0.74 |

Mean: posterior mean; Lower: lower bound of the 95% credible interval; Upper: upper bound of the 95% credible interval; BC: bent-cable model; RE: shared random effect association structure; PW: piecewise model; CV: current value association structure; FHS: the Framingham Heart Study.

Cognitive z-score trajectories in the memory and language domains are shown in **Fig. S2**. The slope parameters suggested a flat trajectory followed by a drastic decline in cognitive functions for both the memory and language domains. The cognitive trajectories demonstrated a 12-year and 14-year change period for the memory and language domains, respectively. On average, the changepoint was at the age of 86 (*β*_*τ*10_ = 26) for the memory domain and the age of 90 (*β*_*τ*20_ = 30) for the language domain. The random changepoints for both cognitive domains showed a high correlation *ρ*_*τ*_ of 0.9.

For the survival process from the healthy state to the dementia state, women had a smaller risk of developing dementia adjusting for other covariates compared to men (hazard ratio [HR]=0.55, 95%CI: 0.31 to 0.93). A longer duration of education corresponded to a smaller risk for dementia (HR=0.87, 95%CI: 0.78 to 0.96). An increase of one year in the changepoint for the memory domain was associated with a 26% lower hazard (HR=0.74, 95%CI: 0.69 to 0.79) for dementia. Of note, the effect of the random intercept of the memory cognitive domain was negative but not significant (HR=0.73, 95% CI: 0.36 to 1.52).

For the survival process from the healthy state to the death state, women had a lower risk of death compared to men (HR=0.55, 95% CI: 0.4 to 0.77). Participants with later changepoint occurrence for the memory domain were less likely to die compared to those with earlier changepoint occurrence in the participants without dementia (HR=0.92, 95% CI: 0.89 to 0.96).

For the transition from dementia to death, we observed that having a longer period of education (HR=1.13, 95% CI:1.01 to 1.26) and a later occurrence of changepoint (HR=1.06, 95% CI:1.02 to 1.11) increased the risk of death for the participants with dementia. It was expected because cognitive reserve for individuals with higher education contributes to more severe underlying brain lesions at the onset of dementia, resulting in a shorter subsequent survival period according to previous studies [36, 37]. Besides, those with later changepoint occurrence might develop dementia at an older age, which could lead to a faster progression to death after the onset of dementia. The effect of changepoint might change if we further include the age at dementia onset as a covariate or use the Markovian model in the conditional hazard function in the illness-death model [17]. For model comparison, most of the findings in the simulation section applied to our real data.

## 7 Discussion

In this paper, we propose a novel joint model that incorporates a random changepoint model for longitudinal cognitive decline and an illness-death model for semi-competing risks. We demonstrate that the proposed joint model, especially the model with a smooth longitudinal change and a correctly specified association structure, accurately estimates the longitudinal changepoint and the impact of longitudinal measures on health state transitions across different data structures. In addition, we compare two changepoint formulations and two association structures in two cohort types. We show that the estimation of longitudinal parameters in the joint model may be influenced by the selection of changepoint formulations and association structures. In contrast, the survival parameter estimates prove more robust and effectively capture the relationship between the longitudinal and survival processes even with misspecification. Compared to the disease cohort, the improvement of the model with a smooth change over a more parsimonious model with an abrupt change is diminished in the community cohort, where participants have less frequent visits and low event rates.

The application in the FHS Offspring cohort suggests that the FHS data may resemble a structure where longitudinal cognitive decline displays a smooth change over time, linked with times to dementia and death by the random changepoint. Additionally, we show that sex, education, and the timing of changepoints are associated with transitions between different health states.

The findings in our study are consistent with prior research. First, previous changepoint joint models demonstrate that changepoints estimated from cognitive scores are associated with the risk for dementia or death [9, 13, 14]. These associations correspond with the results in our study. Second, Yang et al. (2013) observe that the model with an additional smooth interval allows the identification of changepoints for cognitive decline in a later time window compared to the broken-stick model [8], which is in line with our results regarding model comparison. Additionally, Wang (2021) compares various formulations of changepoints within the joint model framework [13]. This work suggests that both the bent-cable model and the smooth polynomial model could effectively characterize the data with a changepoint in a disease cohort, which aligns with our results.

Our study has some limitations. First, it is assumed that all participants will eventually experience a changepoint in their cognitive scores and develop dementia if they are followed long enough. To eliminate this assumption, the cure rate model that allows a fraction of participants to have a null risk of developing events of interest has been suggested in previous joint model literature [10, 38]. Second, a participant may develop dementia between his/her dementia diagnosis date and the preceding visit. Our method considers the time to dementia onset as the time at the dementia diagnosis date, potentially introducing bias due to interval censoring. Third, an FHS participant may be required to undergo additional NP testing annually on condition that his/her test score drops significantly or falls below an education-adjusted cutoff, making the visitation process informative. Although this study design contributes to more frequent observations, failing to account for the informative visitation process may result in biased longitudinal estimates [39]. Plourde (2020) proposes a joint model by treating the visitation process as repeated events using the FHS data [39].

Our method can be extended in the following ways. First, the method can be modified to account for interval censoring in the survival sub-model. Approaches to handle interval censoring have been used in the illness-death models and joint models [40]. Rouanet et al. (2016) handles both interval censoring and semi-competing risks in a joint model framework, but the correlation between cognitive measures and times to dementia or death is captured by latent classes [15]. Second, a participant may experience mild cognitive impairment (MCI) before progressing to dementia or death. To incorporate additional non-terminal health states such as MCI, the illness-death model can be extended to a multistate model within the joint model framework [41]. Dantan et al. (2010) includes a latent pre-diagnosis health state before dementia onset, which is closely related to MCI. Ferrer et al. (2016) proposes a joint model that combines a linear mixed effects model with a multistate model [42].

In conclusion, we propose a joint model that integrates a multivariate random changepoint model for longitudinal cognitive decline with an illness-death model for semi-competing risks. Our proposed model provides a flexible framework for estimating longitudinal trajectories with changepoints and for characterizing the influence of longitudinal measures on transitions between health states. We demonstrate that the selection of changepoint formulations and association structures influences model performance for different cohort structures. Our method enhances understanding of cognitive health while accounting for the complexities introduced by semi-competing risks.

## Acknowledgements

We acknowledge the Framingham Heart Study participants for their contribution to the data.

## Consent to participate

Informed consent was obtained from all individual participants included in the study.

## Data availability statement

The real data that support the findings of this study can be requested at https://www.ncbi.nlm.nih.gov/projects/gap/cgi-bin/document.cgi?study_id=phs000007.v33.p14&phv=523294&phd=4398&pha=4313&pht=12882&phvf=&phdf=&phaf=&phtf=&dssp=1&consent=&temp=1

