## Supplementary methods, tables, and figures for "Joint Model with Random Changepoints for Longitudinal Measures and Semi-Competing Risks"

**Supplementary Materials**

Xuzhi Wang[1], Yorghos Tripodis[1], Michael LaValley[1], Chunyu Liu[1]

1. Department of Biostatistics, Boston University School of Public Health, Boston, MA, 02118, United States

Address: 715 Albany Street, Boston, MA 02118

**A Simulation Steps for Semi-competing Risks**

We followed the cause-specific hazard approach which was described in Beyersmann et al. (2009)[1] and Bender et al. (2005)[2] to generate the time-to-event endpoints. Specifically, this approach is divided into the following several steps:

**Step 1**. Let $S_1(t) = \exp(-H_1(t) - H_2(t))$, where $H_1(t)$ and $H_2(t)$ are the cumulative hazard functions for equations in the illness-death model. Then we simulated a variable $u_i \sim Uniform\ (0,1)$. Next, we obtained $T_i^*$ by solving for the equation $u_i = S_1(t)$. $T_i^*$ was the time to the first event, i.e., dementia or death without dementia.

**Step 2**. We ran a Bernoulli experiment for $T_i^*$ to decide which event occurred at time $T_{\mathrm{i}}^*$ . Specifically, $u_i^*$ was generated from $Uniform\ (0,1)$. If $u_i^* \leq \frac{h_1(T_i^*)}{h_1(T_i^*)+h_2(T_i^*)}$, $T_i^*$ was denoted as the time to dementia, i.e., $T_i^* = T_{1i}$. Otherwise, $T_i^*$ was considered as the time to death without dementia, i.e., $T_i^* = T_{2i}$.

**Step 3**. If $T_i^* = T_{1i}$, which indicated a participant developed dementia at time $T_{1i}$, we further simulated $T_{12i}$ based on the hazard function $h_3(t)$. We then defined $C_i$ as the time to the maximum time of follow-up for each participant. We set the observed outcome information as (time to event 1, time from event 1 to event 2, indicator for dementia, indicator for death), which can be demonstrated in the following scenarios:

- $(T_{1i}, T_{12i}, 1, 1),\ if\ T_{1i} + T_{12i} \leq C_i$
- $(T_{1i}, C_i - T_{1i}, 1, 0),\ if\ T_{1i} + T_{12i} > C_i\ \&\ T_{1i} \leq C_i$
- $(T_{2i}, 0, 0, 1),\ if\ T_{2i} \leq C_i$
- $(C_i, 0, 0,0),\ if\ T_{1i} > C_i\ or\ T_{2i} > C_i$

(1)

### B Supplementary Tables

**Table S1. Simulation results for Scenario 2 (disease cohort with current value association structure).**

| | | BC+RE | | | PW+RE | | | BC+CV | | | PW+CV | | |
|---|---|---|---|---|---|---|---|---|---|---|---|---|---|
| Parameter | True | Mean | PB | CP | Mean | PB | CP | Mean | PB | CP | Mean | PB | CP |
| **Longitudinal** | | | | | | | | | | | | | |
| $\beta_{10}$ | -0.18 | -0.18 | -0.61 | 0.96 | -0.18 | -1.21 | 0.95 | -0.18 | -0.41 | 0.97 | -0.18 | -1.17 | 0.97 |
| $\beta_{11}$ | -0.02 | -0.02 | -1.28 | 0.94 | -0.02 | 1.21 | 0.94 | -0.02 | 0.15 | 0.95 | -0.02 | 3.02 | 0.83 |
| $\beta_{12}$ | -0.15 | -0.14 | -5.62 | 0.71 | -0.13 | -10.09 | 0.13 | -0.15 | 1.73 | 0.95 | -0.14 | -4.00 | 0.79 |
| $\beta_{20}$ | -0.03 | -0.03 | 4.05 | 0.92 | -0.03 | -2.61 | 0.93 | -0.03 | 3.70 | 0.91 | -0.03 | -2.10 | 0.92 |
| $\beta_{21}$ | -0.02 | -0.02 | -0.40 | 0.96 | -0.02 | 22.40 | 0.00 | -0.02 | -0.36 | 0.96 | -0.02 | 22.72 | 0.00 |
| $\beta_{22}$ | -0.20 | -0.20 | 0.04 | 0.96 | -0.18 | -11.52 | 0.00 | -0.20 | 0.33 | 0.95 | -0.18 | -11.41 | 0.00 |
| $\gamma_1$ | 2.00 | 1.82 | -9.03 | 0.97 | - | - | - | 2.01 | 0.67 | 0.96 | - | - | - |
| $\gamma_2$ | 4.00 | 4.00 | -0.11 | 0.97 | - | - | - | 4.04 | 0.94 | 0.95 | - | - | - |
| **Changepoint** | | | | | | | | | | | | | |
| $\beta_{\tau 10}$ | 15.00 | 14.90 | -0.65 | 0.93 | 14.68 | -2.13 | 0.67 | 15.08 | 0.56 | 0.95 | 14.85 | -0.98 | 0.85 |
| $\beta_{\tau 20}$ | 12.00 | 11.94 | -0.49 | 0.92 | 11.48 | -4.31 | 0.03 | 12.02 | 0.20 | 0.95 | 11.57 | -3.62 | 0.10 |
| $\beta_{\tau 11}$ | -3.00 | -2.83 | -5.64 | 0.94 | -2.84 | -5.49 | 0.93 | -2.98 | -0.60 | 0.95 | -2.99 | -0.46 | 0.95 |
| $\beta_{\tau 21}$ | -5.00 | -4.96 | -0.70 | 0.91 | -4.82 | -3.56 | 0.90 | -5.01 | 0.24 | 0.91 | -4.87 | -2.66 | 0.92 |
| $\sigma_{\tau 1}$ | 3.00 | 3.28 | 9.42 | 0.52 | 3.26 | 8.55 | 0.55 | 3.05 | 1.60 | 0.93 | 3.06 | 2.08 | 0.94 |
| $\sigma_{\tau 2}$ | 3.00 | 3.00 | -0.15 | 0.97 | 2.96 | -1.41 | 0.93 | 3.00 | 0.14 | 0.97 | 2.97 | -1.13 | 0.93 |
| $\rho_\tau$ | 0.80 | 0.73 | -9.37 | 0.40 | 0.74 | -8.08 | 0.51 | 0.79 | -0.88 | 0.95 | 0.79 | -1.50 | 0.95 |
| **Survival** | | | | | | | | | | | | | |
| $\alpha_1$ | -0.20 | -0.14 | - | - | -0.14 | - | - | -0.20 | 1.01 | 0.93 | -0.20 | 1.12 | 0.94 |
| $\zeta_1$ | -2.50 | -0.28 | - | - | -0.28 | - | - | -2.54 | 1.43 | 0.92 | -2.55 | 2.06 | 0.92 |
| $\alpha_2$ | -0.40 | -0.38 | - | - | -0.38 | - | - | -0.39 | -1.77 | 0.94 | -0.39 | -1.67 | 0.94 |
| $\zeta_2$ | -0.60 | -0.05 | - | - | -0.06 | - | - | -0.58 | -3.85 | 0.94 | -0.58 | -3.19 | 0.93 |
| $\alpha_3$ | -0.20 | -0.16 | - | - | -0.16 | - | - | -0.20 | -1.03 | 0.97 | -0.20 | -1.42 | 0.97 |
| $\zeta_3$ | -0.50 | -0.05 | - | - | -0.05 | - | - | -0.52 | 4.51 | 0.97 | -0.54 | 7.04 | 0.95 |

True, parameter true value; Mean, posterior mean; PB, percent bias; CP, coverage probability; BC, bent-cable model; RE, shared random effect association structure; PW, piecewise model; CV, current value association structure.

**Table S2. Simulation results for Scenario 3 (community cohort with shared random effect association structure).**

| | | BC+RE | | | PW+RE | | | BC+CV | | | PW+CV | | |
|---|---|---|---|---|---|---|---|---|---|---|---|---|---|
| Parameter | True | Mean | PB | CP | Mean | PB | CP | Mean | PB | CP | Mean | PB | CP |
| **Longitudinal** | | | | | | | | | | | | | |
| $\beta_{10}$ | -0.18 | -0.18 | 0.80 | 0.94 | -0.18 | 0.47 | 0.94 | -0.18 | 0.61 | 0.95 | -0.18 | 0.27 | 0.95 |
| $\beta_{11}$ | -0.02 | -0.02 | -0.21 | 0.93 | -0.02 | 0.89 | 0.90 | -0.02 | 1.63 | 0.91 | -0.02 | 3.06 | 0.83 |
| $\beta_{12}$ | -0.15 | -0.17 | 14.28 | 0.94 | -0.15 | -2.45 | 0.96 | -0.19 | 24.75 | 0.74 | -0.16 | 7.28 | 0.89 |
| $\beta_{20}$ | -0.03 | -0.03 | 2.17 | 0.94 | -0.03 | -1.06 | 0.95 | -0.03 | 2.32 | 0.94 | -0.03 | -1.30 | 0.95 |
| $\beta_{21}$ | -0.02 | -0.02 | 0.10 | 0.97 | -0.02 | 2.04 | 0.85 | -0.02 | 0.70 | 0.96 | -0.02 | 2.58 | 0.83 |
| $\beta_{22}$ | -0.20 | -0.21 | 2.86 | 0.95 | -0.16 | -18.38 | 0.20 | -0.20 | 1.12 | 0.96 | -0.16 | -18.54 | 0.17 |
| $\gamma_1$ | 2.00 | 2.96 | 48.10 | 0.98 | - | - | - | 3.31 | 65.25 | 0.98 | - | - | - |
| $\gamma_2$ | 4.00 | 4.16 | 3.96 | 0.96 | - | - | - | 4.10 | 2.42 | 0.96 | - | - | - |
| **Changepoint** | | | | | | | | | | | | | |
| $\beta_{\tau 10}$ | 25.00 | 25.81 | 3.23 | 0.92 | 25.10 | 0.41 | 0.97 | 28.18 | 12.72 | 0.15 | 27.72 | 10.87 | 0.33 |
| $\beta_{\tau 20}$ | 27.00 | 27.40 | 1.49 | 0.96 | 26.03 | -3.61 | 0.82 | 28.43 | 5.31 | 0.67 | 27.46 | 1.70 | 0.92 |
| $\beta_{\tau 11}$ | -3.00 | -2.94 | -1.88 | 0.97 | -3.01 | 0.42 | 0.97 | -3.40 | 13.17 | 0.96 | -3.55 | 18.43 | 0.95 |
| $\beta_{\tau 21}$ | -5.00 | -4.98 | -0.45 | 0.93 | -5.04 | 0.76 | 0.94 | -5.23 | 4.56 | 0.93 | -5.46 | 9.10 | 0.93 |
| $\sigma_{\tau 1}$ | 8.00 | 8.13 | 1.64 | 0.96 | 8.18 | 2.20 | 0.96 | 8.57 | 7.14 | 0.88 | 8.71 | 8.84 | 0.83 |
| $\sigma_{\tau 2}$ | 8.00 | 8.15 | 1.93 | 0.93 | 8.06 | 0.70 | 0.93 | 8.40 | 4.98 | 0.86 | 8.45 | 5.61 | 0.89 |
| $\rho_{\tau}$ | 0.80 | 0.79 | -1.45 | 0.97 | 0.80 | 0.44 | 0.96 | 0.80 | -0.26 | 0.97 | 0.81 | 1.41 | 0.92 |
| **Survival** | | | | | | | | | | | | | |
| $\alpha_1$ | -0.60 | -0.66 | 9.62 | 0.94 | -0.68 | 13.01 | 0.92 | -0.50 | - | - | -0.49 | - | - |
| $\zeta_1$ | -0.30 | -0.32 | 6.37 | 0.93 | -0.32 | 7.04 | 0.92 | -1.53 | - | - | -1.52 | - | - |
| $\alpha_2$ | -0.60 | -0.61 | 2.03 | 0.97 | -0.62 | 3.22 | 0.97 | -0.57 | - | - | -0.57 | - | - |
| $\zeta_2$ | -0.10 | -0.10 | -4.67 | 0.92 | -0.09 | -7.12 | 0.94 | -0.32 | - | - | -0.32 | - | - |
| $\alpha_3$ | -0.10 | -0.15 | 50.11 | 0.97 | -0.16 | 57.49 | 0.97 | -0.08 | - | - | -0.07 | - | - |
| $\zeta_3$ | -0.10 | -0.12 | 16.22 | 0.96 | -0.12 | 15.78 | 0.94 | -0.29 | - | - | -0.28 | - | - |

True, parameter true value; Mean, posterior mean; PB, percent bias; CP, coverage probability; BC, bent-cable model; RE, shared random effect association structure; PW, piecewise model; CV, current value association structure.

**Table S3. Simulation results for Scenario 4 (community cohort with current value association structure).**

| | | BC+RE | | | PW+RE | | | BC+CV | | | PW+CV | | |
|---|---|---|---|---|---|---|---|---|---|---|---|---|---|
| Parameter | True | Mean | PB | CP | Mean | PB | CP | Mean | PB | CP | Mean | PB | CP |
| **Longitudinal** | | | | | | | | | | | | | |
| $\beta_{10}$ | -0.18 | -0.18 | -1.03 | 0.95 | -0.18 | -1.41 | 0.94 | -0.18 | 0.89 | 0.94 | -0.18 | 0.50 | 0.93 |
| $\beta_{11}$ | -0.02 | -0.02 | -6.43 | 0.71 | -0.02 | -4.28 | 0.83 | -0.02 | -0.17 | 0.91 | -0.02 | 1.03 | 0.89 |
| $\beta_{12}$ | -0.15 | -0.13 | -11.91 | 0.77 | -0.12 | -23.02 | 0.38 | -0.17 | 13.87 | 0.89 | -0.15 | -2.90 | 0.90 |
| $\beta_{20}$ | -0.03 | -0.03 | 0.20 | 0.95 | -0.03 | -1.45 | 0.95 | -0.03 | 0.62 | 0.94 | -0.03 | -2.70 | 0.93 |
| $\beta_{21}$ | -0.02 | -0.02 | 0.00 | 0.95 | -0.02 | 2.10 | 0.88 | -0.02 | 0.33 | 0.93 | -0.02 | 2.27 | 0.87 |
| $\beta_{22}$ | -0.20 | -0.21 | 2.83 | 0.91 | -0.17 | -16.63 | 0.27 | -0.20 | 2.08 | 0.94 | -0.17 | -17.02 | 0.23 |
| $\gamma_1$ | 2.00 | 3.89 | 94.63 | 0.95 | - | - | - | 3.17 | 58.59 | 0.98 | - | - | - |
| $\gamma_2$ | 4.00 | 4.04 | 0.94 | 0.97 | - | - | - | 4.04 | 0.99 | 0.96 | - | - | - |
| **Changepoint** | | | | | | | | | | | | | |
| $\beta_{\tau 10}$ | 25.00 | 24.10 | -3.59 | 0.89 | 23.47 | -6.13 | 0.70 | 26.08 | 4.31 | 0.93 | 25.34 | 1.35 | 0.94 |
| $\beta_{\tau 20}$ | 27.00 | 26.21 | -2.94 | 0.92 | 24.92 | -7.70 | 0.45 | 27.26 | 0.97 | 0.99 | 26.01 | -3.66 | 0.84 |
| $\beta_{\tau 11}$ | -3.00 | -2.41 | -19.52 | 0.91 | -2.46 | -18.14 | 0.93 | -3.15 | 5.15 | 0.95 | -3.09 | 3.06 | 0.97 |
| $\beta_{\tau 21}$ | -5.00 | -4.61 | -7.83 | 0.93 | -4.65 | -7.08 | 0.95 | -5.02 | 0.47 | 0.94 | -4.99 | -0.21 | 0.95 |
| $\sigma_{\tau 1}$ | 8.00 | 9.17 | 14.59 | 0.59 | 9.18 | 14.76 | 0.59 | 8.26 | 3.31 | 0.92 | 8.28 | 3.46 | 0.91 |
| $\sigma_{\tau 2}$ | 8.00 | 7.91 | -1.15 | 0.95 | 7.81 | -2.33 | 0.93 | 8.09 | 1.17 | 0.94 | 7.98 | -0.30 | 0.96 |
| $\rho_\tau$ | 0.80 | 0.74 | -7.90 | 0.72 | 0.76 | -5.41 | 0.87 | 0.79 | -1.84 | 0.95 | 0.80 | -0.26 | 0.96 |
| **Survival** | | | | | | | | | | | | | |
| $\alpha_1$ | -0.20 | -0.15 | - | - | -0.15 | - | - | -0.19 | -5.34 | 0.93 | -0.19 | -5.88 | 0.92 |
| $\zeta_1$ | -2.50 | -0.21 | - | - | -0.20 | - | - | -2.55 | 1.87 | 0.92 | -2.55 | 2.07 | 0.90 |
| $\alpha_2$ | -0.40 | -0.38 | - | - | -0.39 | - | - | -0.39 | -2.22 | 0.96 | -0.40 | -0.53 | 0.96 |
| $\zeta_2$ | -0.60 | -0.01 | - | - | -0.01 | - | - | -0.59 | -2.29 | 0.92 | -0.58 | -2.98 | 0.92 |
| $\alpha_3$ | -0.20 | -0.19 | - | - | -0.19 | - | - | -0.22 | 12.19 | 0.91 | -0.22 | 9.02 | 0.90 |
| $\zeta_3$ | -0.50 | -0.03 | - | - | -0.03 | - | - | -0.56 | 11.23 | 0.95 | -0.57 | 14.47 | 0.95 |

True, parameter true value; Mean, posterior mean; PB, percent bias; CP, coverage probability; BC, bent-cable model; RE, shared random effect association structure; PW, piecewise model; CV, current value association structure.

**Table S4. Mean WAIC values for each model in each scenario.**

| | Scenario 1 | Scenario 2 | Scenario 3 | Scenario 4 |
|---|---|---|---|---|
| BC+RE | **7616** | 7904 | 4113 | 5559 |
| PW+RE | 7716 | 8004 | **4108** | 5557 |
| BC+CV | 7777 | **7536** | 4363 | 5522 |
| PW+CV | 7880 | 7639 | 4367 | **5509** |

BC, bent-cable model; RE, shared random effect association structure; PW, piecewise model; CV, current value association structure.

**Table S5. Neuropsychological tests included in the memory and language domains.**

| Domains | Tests |
|---|---|
| Memory | Wechsler Memory Scale (WMS) Logical Memory – Immediate Recall;<br>WMS Logical Memory – Delayed Recall;<br>WMS Visual Reproductions – Immediate Recall;<br>WMS Visual Reproductions – Delayed Recall;<br>WMS Paired Associates – Immediate Recall;<br>WMS Paired Associates – Delayed Recall |
| Language | Boston Naming Test 30 item version;<br>Wide Range Achievement Test-3 Reading subtest;<br>Wechsler Adult Intelligence Scale Similarities subtest |

### C Supplementary Figures

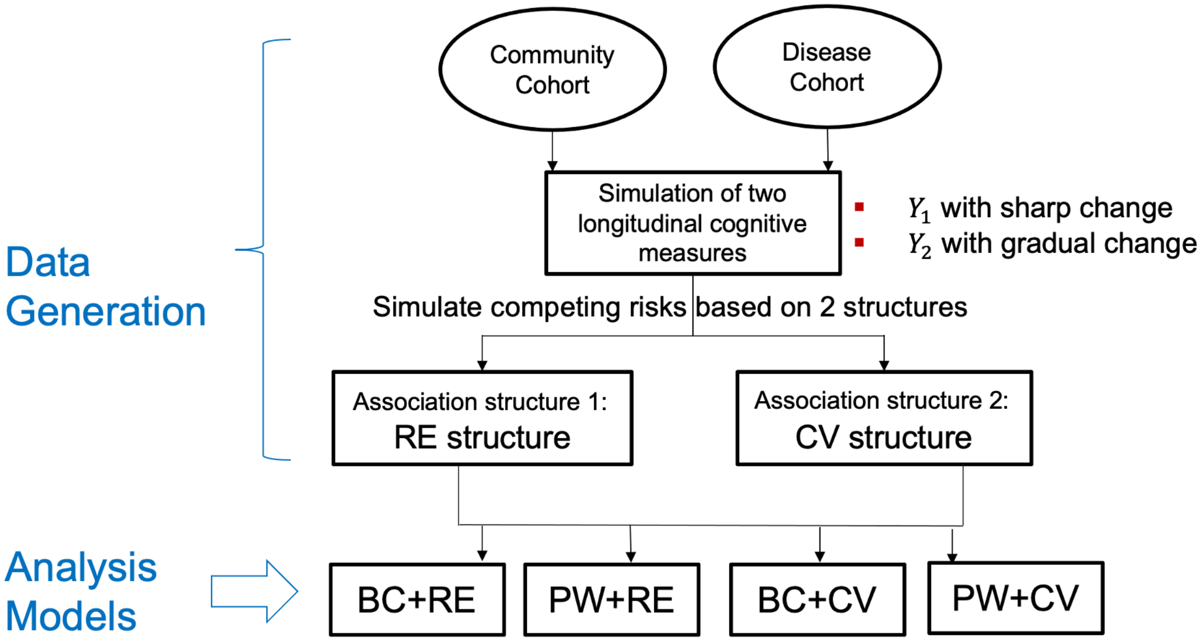


**Fig. S1** Flowchart of the simulation study. Four simulation scenarios are generated: disease cohort with RE structure, disease cohort with CV structure, community cohort with RE structure, and community cohort with CV structure. BC, bent-cable model; RE, shared random effect association structure; PW, piecewise model; CV, current value association structure

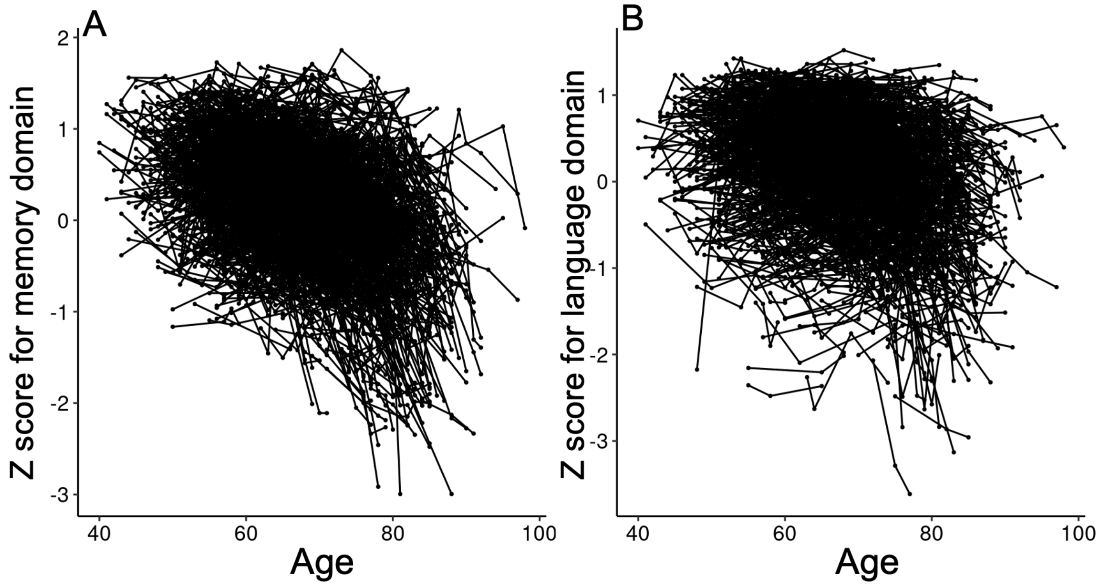


**Fig. S2** Z-score trajectories for the memory and language domains of neuropsychological tests in the FHS Offspring cohort. Panels A and B show individual z-score trajectories for the memory and language domains, respectively. In each panel, the x-axis represents age, and the y-axis represents z-scores. Each curve denotes a participant's trajectory. FHS: Framingham Heart Study.
